# A comprehensive tandem repeat catalog of the human genome

**DOI:** 10.1101/2024.06.19.24309173

**Authors:** Readman Chiu, Indhu-Shree Rajan-Babu, Jan M. Friedman, Inanc Birol

## Abstract

With the increasing availability of long-read sequencing data, high-quality human genome assemblies, and software for fully characterizing tandem repeats, genome-wide genotyping of tandem repeat loci on a population scale becomes more feasible. Such efforts not only expand our knowledge of the tandem repeat landscape in the human genome but also enhance our ability to differentiate pathogenic tandem repeat mutations from benign polymorphisms. To this end, we analyzed 272 genomes assembled using datasets from three public initiatives that employed different long-read sequencing technologies. Here, we report a catalog of over 18 million tandem repeat loci, many of which were previously unannotated. Some of these loci are highly polymorphic, and many of them reside within coding sequences.

## Introduction

Tandem repeats (TRs), including simple sequence repeats or short tandem repeats (STRs) with 1 to 6 base pair (bp) repeat motifs^1^ and variable number tandem repeats (VNTRs) with >7 bp repeat motifs^2^, constitute about 7 to 8% of the human genome^3,4^. Many TR loci are highly polymorphic and exhibit marked divergence in length and sequence composition (i.e., the number and types of repeat motifs they harbor)^2,5,6^. These variations, particularly the changes in repeat length or “expansions” over a specific threshold, cause more than 65 neurological and 14 neuromuscular disorders, with pathogenic expansions at some loci ranging several kilobases (kb) in length^6–9^. The length and sequence composition of the expanded TR allele can significantly influence the age of onset, penetrance, severity, and/or clinical presentation in patients with some repeat expansion disorders^6^. Recently, studies have also revealed associations between variations in TRs and cancer, autism, and other complex traits and disorders^10–16^.

Over 2 million TRs have been reported to occur in the human genome^17,18^, residing within both coding and non-coding regions and playing a crucial role in regulating gene expression, genome stability, and chromatin structure^19–21^. Yet, TR expansions have long been disregarded as a possible explanation of unexplained genetic disorders that are not neurological, largely due to the challenges in profiling them in high-throughput genomic sequencing datasets and in interpreting their clinical relevance^22^. Consequently, our understanding of the extent of variability within these loci and their contribution to disease, beyond the catalog of known disease-associated TRs^8,9,23^, are quite limited. Although computational methods are now available to detect repeat expansions in next-generation sequencing (NGS) data^24–29^, the inherent read-length limitation of these technologies means that only TRs encompassed by sequencing reads (∼150 to 250 base pairs (bp) in length) can be accurately genotyped and characterized using NGS^30^. These challenges are now being overcome with the adoption of Oxford Nanopore Technologies (ONT) and Pacific Biosciences (PacBio) long-read sequencing (LRS) approaches, which yield reads that are ≥15 to 100 kb in length. In recent years, LRS has facilitated the discovery of several novel disease-associated TR expansions^31^ by offering an unprecedented advantage over NGS and low-throughput clinical repeat-primed polymerase chain reaction and Southern blot assays in determining the length, sequence composition, and epigenetic signatures across genome-wide TRs of diverse sizes^6,32^.

Differentiating pathogenic TR expansions from benign polymorphic variations requires prior knowledge of the range of repeat length variability of each TR locus in the general population. While population databases such as the Genome Aggregation Database and the Database of Genomic Variants have been fundamental to the prioritization and identification of rare pathogenic sequence and copy number variations^33,34^, there has been a paucity of TR variation databases derived from LRS datasets^32^, which are more accurate in revealing the genotypes of TRs greater than ∼300 to 500 bp in size. A comprehensive and accurate catalog of variations of all TRs genome-wide built from LRS would facilitate the discovery of novel disease-associated TR loci and may help account for some of the missing heritability among patients with undiagnosed rare and complex disorders.

In addition to improving variant calling, LRS has also made the construction of the telomere-to-telomere sequence of the hydatidiform mole (T2T-CHM13)^35^ and high-quality haplotype-resolved assemblies of human genomes^36^ possible in recent times. Utilization of T2T-CHM13 as a reference has been shown to markedly improve structural variant calling and analysis of genomic regions that remain unresolved in the GRCh38 reference sequence^37^, but if and to what extent the T2T-CHM13 reference can improve TR calling compared to GRCh38 remains unclear. Moreover, the availability of haplotype-resolved assemblies allows us now to detect and annotate TRs *de novo* (i.e., genotype and characterize TRs directly from the assembled contigs), mitigating some of the errors and biases associated with alignment-based variant calling and allowing for direct comparison and complementation of alignment-based TR calls with those of assembly-based TR calls.

In this study, we leverage the strengths of LRS data, combined with alignments to GRCh38 and T2T-CHM13 references as well as haplotype-resolved assemblies, in creating the most comprehensive TR catalog available to date and outlining the best approach to curate TRs. We have cataloged the genotypes of 18 million TRs in ONT and/or PacBio HiFi whole-genome LRS data from a total of 272 individuals of diverse ancestries sequenced by the Human Pangenome Reference Consortium (HPRC)^36^, the Human Genomes Structural Variation Consortium (HGSVC, phase 2 or HGSVC2)^38^, or the 1000 Genomes ONT Sequencing Consortium (1KGP-ONT)^39^.

## Results

### Data

The source of LRS data used to generate our catalog (Fig. 1a) originates from: 1) The year one data freeze of HPRC which consists of 47 1KGP samples with haplotype-resolved Hifiasm^40^ assemblies constructed with PacBio HiFi data and short-read data on both parents of each individual (trio Hifiasm). Thirty-nine of these assemblies were augmented with ONT PromethION sequence data. More recently, HPRC released PacBio HiFi data from an additional 91 samples. We assembled these, and five year-one-data-freeze samples without any assembled sequence releases, using Hifiasm in default mode because we could not locate the parent sequence data; 2) The HGSVC2 release consisting of 35 1KGP samples sequenced with PacBio Continuous Long Reads (CLR) or HiFi sequencing. We included 30 of them that are not part of the HPRC dataset. The CLR data were assembled with Flye^41^ and the HiFi data were assembled with Racon^42^, respectively; and 3) The first public release of the 1KPG-ONT dataset consisting of ONT sequencing data with diploid assemblies generated using the Shasta+Hapdup pipeline^43^. We included 99 of the 100 1KGP sequenced samples to our cohort, omitting one (HG02615) that had already been sequenced by the HPRC consortium. The online locations of the three data sources and a detailed account of data type per sample are provided in Supplementary Tables 1 and 2, respectively.

**Figure 1.**
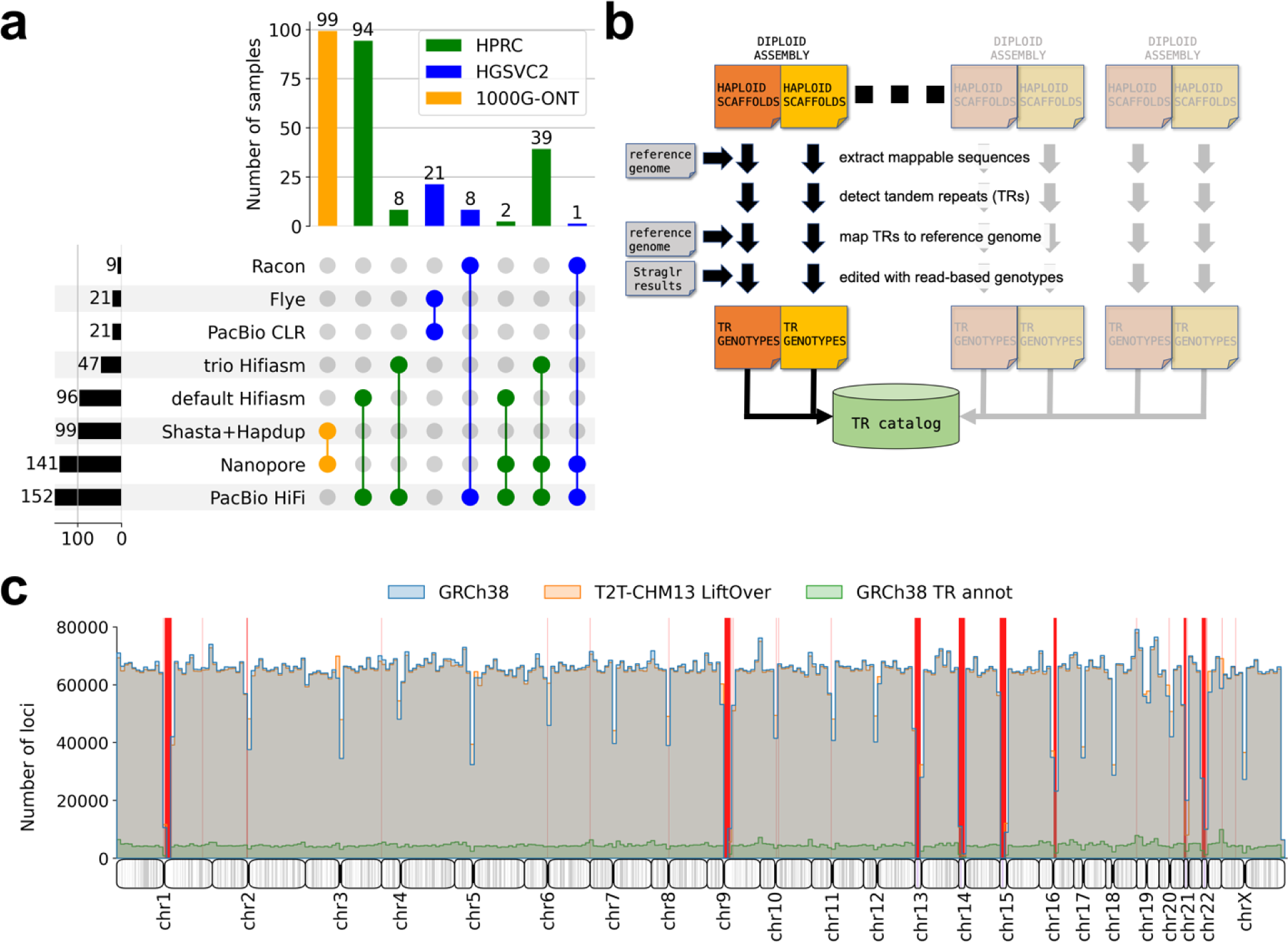
**a.** Data types and sources of long-read samples used in this study. **b.** Pipeline design to capture TRs from diploid long-read assemblies. **c.** Histogram showing the distribution of TR loci genotyped in the GRCh38 catalog (blue) across the human genome (bin size = 1Mb). Distributions of loci from the T2T-CHM13 catalog liftovered to GRCh38 reference coordinates (orange) and the UCSC TR (Simple Repeats + RepeatFinder + Low Complexity) annotations (green) are also shown for comparison (overlapping blue and orange bars lead to grey colors). Red bars indicate GRCh38 sequence gaps.

### *De novo* TR detection

High-quality *de novo* genome assemblies allow us to detect TRs genome-wide unconstrained by previous repeat annotations. Our analysis pipeline (Fig. 1b) starts by extracting TRs from the assemblies, followed by the determination of their genomic locations so that alleles collected from different samples can be grouped and consolidated into a single catalog. We ran our pipeline against both the GRCh38 and T2T-CHM13^44^ references independently, as the latter is gaining wider adoption in the research community as the reference-of-choice for variant calling. We used Tandem Repeat Finder (TRF)^45^ to detect TRs in the generated assemblies owing to its speed and accuracy.

Because TRF processing in heavily TR-laden genomic regions such as the centromeric sequences is computationally infeasible, we began our analysis by first mapping each haplotype assembly to the reference genome (GRCh38 or T2T-CHM13) to identify potentially problematic sequences that gave low-quality or ambiguous mappings. This filtering step significantly boosted TRF’s processing speed on the remaining scaffold sub-sequences, with each of these concurrently executed tasks completing within seconds. TRF was run with a lenient parameter setting (see Method) to incorporate long VNTRs in the genome that often exhibit higher motif heterogeneity. The chosen parameter settings simultaneously led to the inclusion of more short STRs (2-3 copies) with impure repeats than with the default settings. To reduce redundancy we removed TRs entirely subsumed in overlapping neighbours. Flanking sequences of each TR were extracted from the assembly and mapped back to the reference genome to determine their genomic locations. On average, TRF identified between 14.0 and 14.2 million 2-100 bp TRs from the assemblies after initial filtering, and we were able to determine 98.5% and 98.2% of their genomic locations in GRCh38 and T2T-CHM13, respectively.

Concomitantly, HiFi and ONT sequencing reads were aligned against the reference genomes, and the resulting alignments were analysed with Straglr^46^ to scan for expansions that are at least 100 bp greater than the reference. The Straglr results enable cross-checking against the assembly-based genotypes, albeit limited to loci that are “expanded” (∼5,025 and ∼3,004 per sample on average for GRCh38 and T2T-CHM13, respectively) relative to the reference genomes. We expected that TRs with modest deviations (<100 bp) from the reference genome should be reliably captured in the high-quality assemblies, and our analysis below confirmed so. We used high-confidence Straglr results (see Methods for criteria) to edit our assembly-based results in three operations: 1) Replace size-discordant (difference >10%) alleles; 2) Fill in missing haplotypes; and 3) Add loci not captured from the assembly-based pipeline. On average 305 edits (operations 1 and 2) were made, and 105 loci (operation 3) were added to complement the assembly-based genotypes of each sample. We observed that editing was frequently performed on homozygous loci in the assemblies with differing heterozygous genotypes from read-based Straglr results.

When the number of edits and novel loci were broken down by sequencing platform and assembler, we noted that the 21 PacBio CLR samples of the HGSVC2 had substantially more edits performed than the PacBio HiFi and 1KGP ONT samples (Supplementary Table 3). Comparing the repeat size differences between the assembly-based and the read-based genotyped alleles of the edited loci shows that the read-based genotypes in these CLR samples tend to be smaller than the assembly-based genotypes, contrary to what is observed for the HiFi samples (Supplementary Fig. 1, left). Moreover, the sizes of the novel loci introduced into the catalog also tend to be smaller for the CLR samples than the other samples (Supplementary Fig. 1, right). It can be safely assumed that the more erroneous CLR sequencing reads may explain the higher variability in the read-based genotypes and more disagreements from the assembly. However, the fraction of alleles edited out of the total number of over 10 million loci genotyped is so small that it should not substantially affect the quality of the final catalog. On the other hand, since the 1KGP ONT samples have no accompanying PacBio sequencing data, the disagreement between the assembly-based and read-based results is naturally smaller than the HiFi samples, of which ∼30% have orthogonal ONT Straglr results. Many fewer edits were performed in the T2T-CHM13 catalog (Supplementary Table 3), suggesting that the T2T-CHM13 reference captures more representative TR genotypes in the population than the GRCh38 reference.

As the last polishing step, we compared all the individual TR mappings with their parent scaffold mappings done at the beginning of our pipeline and removed any TRs with inconsistent results. Finally, we combined TR alleles from all haplotypes by performing a graph-based clustering of loci with similar genomic coordinates. The most frequently observed motif was used as the consensus motif for a given locus and the repeat copies for each sample were re-calculated based on the length of the consensus motif.

### Catalogs

A total of 18,759,399 loci (13,394,896 STRs and 5,364,503 VNTRs) with 2-100 bp repeated (2 ≥ copies) motifs on chromosomes 1-22 and X have genotype results in the GRCh38 catalog. We did not include any chrY TRs as the mappings of their flanking sequences indicate most of them reside in segmental duplications. Out of these, 16,204,012 (86.4%) loci have at least one haplotype genotyped in at least half (136) of the total number of samples, and 12,966,161 (69.1%) loci have at least half of the maximum number of alleles genotyped (Supplementary Fig. 2). A small degree of redundancy exists where 1,383,557 loci (7.4%) are nested within another locus, typically corresponding to cases of overlapping STRs and VNTRs. The average density of TRs is around 6,163 per 1Mb, with precipitous drops around centromeres and sequence gaps in the GRCh38 reference genome (Fig. 1c). Processing the same 1KGP LRS data with the same pipeline against the T2T-CHM13 reference resulted in a catalog of 19,325,220 loci (13,817,036 STRs and 5,508,184 VNTRs), with only 565,821 loci (3.0%) more than the GRCh38 catalog.

Even though the T2T-CHM13 reference resolves the sequence gaps in GRCh38 that correspond to highly repetitive sequences, there is only marginal gain in the number of TR loci genotyped in the T2T-CHM13 catalog due to the difficulty in mapping TR origins using their flanking sequences, which themselves are likely embedded within highly repetitive regions (e.g., 9q12 highlighted in red in Supplementary Fig. 3). The T2T-CHM13 catalog consists of 16,406,796 (84.9%) loci with at least one haplotype genotyped in at least half of the total number of samples, and 13,103,532 (67.8%) loci with at least half of the maximum number of alleles genotyped. Similar to the GRCh38 catalog, a small subset of loci, 1,584,679 (8.2%), are nested within bigger loci and the average TR density is around 6,297 loci per 1Mb.

Our catalogs contain considerably more TRs than the current standard UCSC TR annotations (Simple Repeats, RepeatFinder, and Low Complexity) as we included shorter and impure TRs. Few exceptions to this relative abundance were observed with the T2T-CHM13 annotation at locations such as 1q12, 12p12, 13p12, 15p11.2-12, 16q11.2, which correspond to satellite DNA regions that are fully sequenced and annotated in the T2T-CHM13 reference genome but not in GRCh38. These regions are beyond the capability of our TR genotyping methodology. Using the UCSC liftOver utility^47^, 18,037,065 loci, representing 96.1% and 93.3%, respectively, of the GRCh38 and T2T-CHM13 catalogs, could be successfully converted directly from one reference to the other and vice versa.

We observed that in a small number of loci, the genotypes from all studied samples consistently agreed with one reference sequence but not the other, suggesting potential sizing errors in one of the references (unless the reference captured a rare allele not seen in our studied samples). To identify these loci, we extracted TRs from each catalog in turn that (a) can be successfully lifted over so that their corresponding sizes in the other reference can be determined; (b) have at least 100 genotype calls across all samples; (c) have a genotype that is at least 100 bp different from the reference genome that the catalog was built on (i.e. potential error); and (d) have a standard deviation (SD) of at least 100 bp indicating limited variability in the genotypes observed for all samples within the catalog. By comparing the mean repeat size from the alleles in the catalog against the TR length in the liftOver reference, we inferred that the liftOver reference is more accurate in sizing a given TR locus if their size difference is within one SD of each other. This exercise identified 1,223 loci where the genotyped repeat sizes consistently agreed with T2T-CHM13 but not GRCh38, and 162 loci where the repeat sizes consistently agreed with GRCh38 but not T2T-CHM13 (Supplementary Table 4).

We annotated both catalog versions with the genomic locations of the genotyped TR loci (see Methods). Approximately 45% of the loci reside within a gene or promoter (defined as 1-1000 bp upstream of a gene), with the majority (∼90%) of them localized in introns and the remaining distributed within untranslated regions (UTRs), coding sequences (CDS), exons, promoters, and exon and transcript boundaries (Table 1).

**Table 1.**
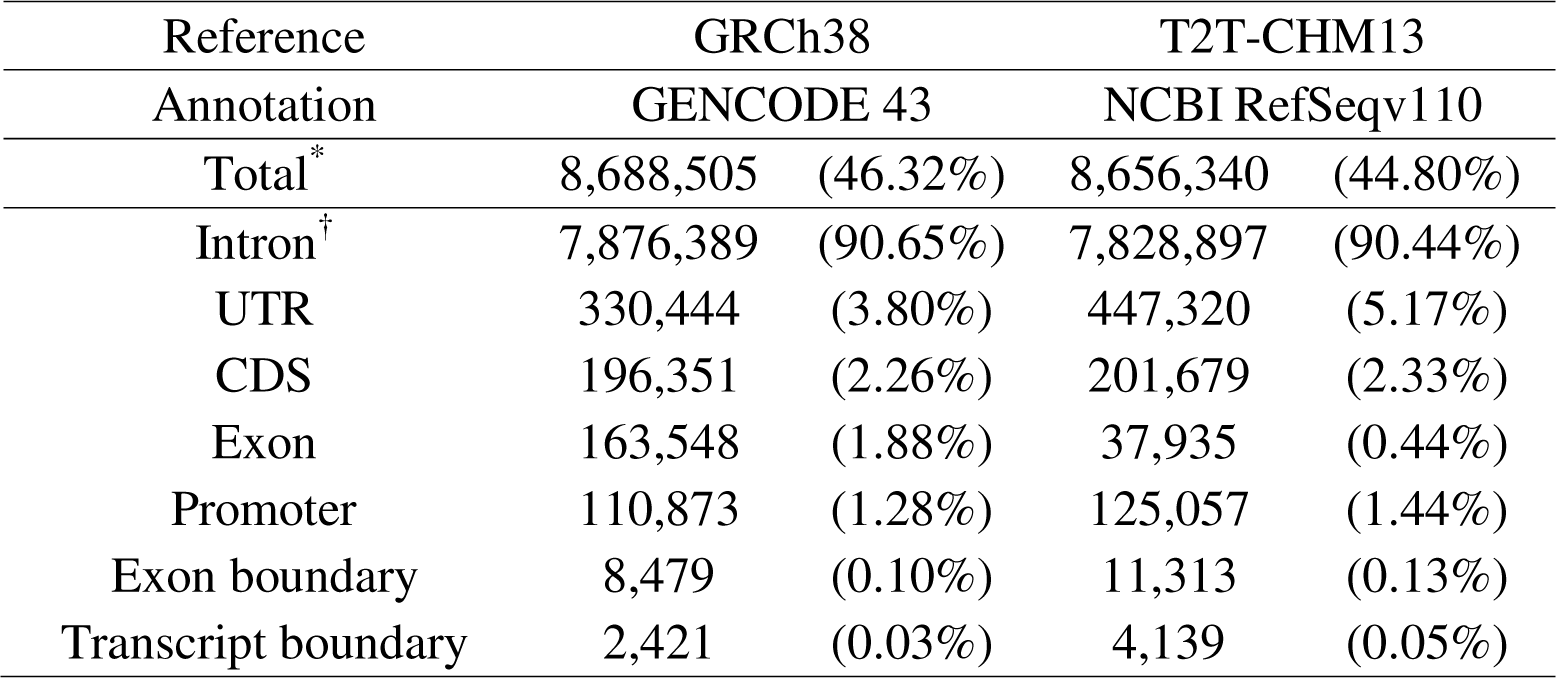
Number of loci located in genic features in both GRCh38 and T2T-CHM13 catalogs.

### Assembly-based vs read-based genotypes

We compared assembly-based genotypes against read-based genotypes (Straglr results on read alignments) from 43 samples that have Nanopore in addition to PacBio HiFi sequencing data to understand how these two sequencing platforms differ from, or complement, each other in TR detection and genotyping. We used the genotype calls from our assembly-based pipeline as the comparison baseline and intersected their coordinates against Straglr results from ONT or PacBio HiFi reads individually. Only alleles that are larger than the GRCh38 reference by at least 100 bp were compared to match the parameter conditions used with Straglr. This resulted in 327,147 assembly-based TR alleles originating from 24,268 loci. In total, 68.4% (223,911) have matching alleles (location spans with at least 80% reciprocal overlaps and sizes within 10% of each other) in both HiFi and ONT Straglr results, 15.1% (49,428) have matching alleles only in HiFi Straglr results, 10.2% (33,536) have matching alleles only in Nanopore Straglr results, and 6.2% (20,299) do not have matching alleles in either HiFi or Nanopore Straglr results (Supplementary Fig. 4, bottom). Comparing the allele size distributions between the four groups (Supplementary Fig. 4, top), alleles from the assembly-Nanopore/Straglr matches contained substantially more alleles at least 10kb in length (413, many of them located in subtelomeric TR arrays) than from the assembly-HiFi/Straglr matches (20), suggesting ONT sequencing reads in these datasets can capture longer TR alleles than HiFi reads overall.

Interestingly, we also found that assemblies (41 Hifiasm + 2 Racon) using HiFi reads were able to reconstruct long TR alleles that were otherwise undetected from genotyping directly on the reads alone, and the high percentage (576/657 = 87.7%) of matching alleles 10kb between ≥ assembly and ONT Straglr results validates the assemblies of these long TR alleles. In fact, two alleles longer than110kb were detected solely from Hifiasm assemblies, unsurprising because they are likely beyond the size range of ONT sequencing reads employed on these samples.

These two alleles originate from a highly polymorphic intergenic 29-bp VNTR on 1q42.2 (chr1:236,097,103-236,097,409) in two different samples, each being heterozygous with a shorter 4kb or 6kb allele, respectively, reconstructed for the second haplotype. These two shorter alleles were also confirmed by Straglr genotyping on the input HiFi reads. The fact that more than half of the sequencing reads mapped to this locus failed to span the entire locus according to Straglr corroborates with the existence of a very long second allele.

Two HPRC samples with Nanopore sequencing data available were not released with trio Hifiasm assemblies. This allowed us to evaluate genotyping results based on default Hifiasm, which was used to assemble 94 HPRC samples. Specifically, we inspected whether default Hifiasm was able to reconstruct large TRs without any supplementary Hi-C^48^ or parental sequencing data. We identified an expanded locus reported by Straglr that is also assembled by default Hifiasm: a VNTR composed of 34-bp repeat units in *TMEM242* on 6q25.3 (chr6:157,310,356-157,314,362) in both samples. From the assembly, heterozygous genotypes, 40.0 kb | 32.8 kb and 38.5 kb | 31.1kb, were called in the HG01442 and HG03471 samples, respectively. Straglr reported 40.0 kb | 32.8 kb and 38.6 kb | 31.6 kb on this locus for the same samples, respectively, based on Nanopore-read alignments. The high level of agreement confirms the ability of default Hifiasm to reconstruct large TR alleles without orthogonal data.

### STR size polymorphism

Of all STRs in the GRCh38 catalog 18.2% (2,286,309) have more than one size genotype across all samples. To measure the extent of size polymorphism, we calculated both standard deviation (SD) and interquartile range (IQR) in repeat copies for every locus in this set. Some loci yielded a high level of variability according to both statistical measures when only two alleles of vastly different lengths were detected across all samples. However, multiple alleles may exist for the same locus despite their relatively small size differences. We attempted to estimate the number of “major” common alleles for a polymorphic locus by clustering alleles to account for potential genotyping errors. Applying a simple clustering of allele sizes based on a relative fractional distance (reciprocal 5% threshold) on STRs with more than one size detected across all samples produced a single cluster for 2,154,430 (94.2%) loci, two clusters for 117,262 (5.1%) loci, 3-12 clusters for 12,521 (0.5%) loci, and zero clusters (<10 alleles for any cluster) for 2,096 (<0.1%) loci. In total, 98.9% (12,418,447) of STRs studied exhibited only one principal size (identical sizes or single cluster) in the studied samples.

A bivariate histogram (Supplementary Fig. 5) showing the counts of loci with varying combinations of IQR and number of clusters demonstrated that the overwhelming majority of STRs genotyped in our samples are non-polymorphic. We also provide visual verification of clustering results in loci demonstrating either high or low IQR (Supplementary Figs. 6-7). Loci exhibiting high IQR but only a small number of clusters (top-left in the bivariate histogram and multiple cases in Supplementary Fig. 6) highlight the existence of highly polymorphic loci for which clustering fails to convey the extent of polymorphism. In total, 1,743 STRs with 100 ≥ distinctive allele sizes genotyped regardless of IQR magnitude or clustering results likely represented the most polymorphic STRs in our study cohort. On the other hand, clustering was successful even at loci with alleles exhibiting a relatively small statistical spread in repeat sizes, suggesting the small variations represent real polymorphisms instead of genotyping errors (Supplementary Fig. 7). An example of this is a GC-rich locus near the splice donor site of exon 3 of *BEGAIN* at chr14:100,546,328-100,546,496. Twelve major alleles for this locus, each observed in at least ten haplotypes within a size range of ∼200bp, were identified by clustering (Fig. 2a).

**Figure 2.**
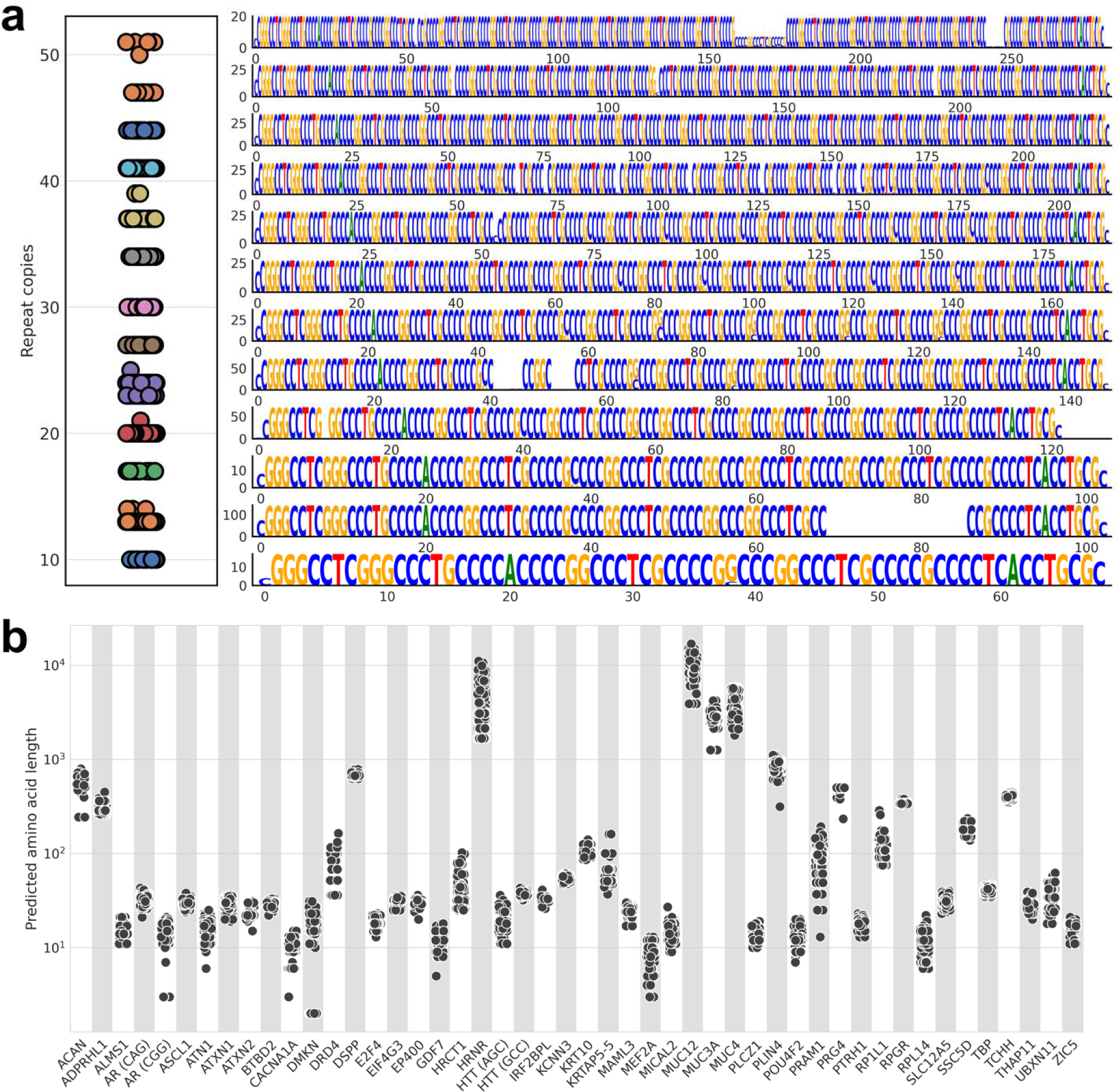
**a.** Size (repeat copies) clustering (left) and sequence logos generated from alleles within each size cluster (right) for the TR locus in *BEGAIN* (chr14:100,546,328-100,546,496). Dots of the same color (left) represent alleles segregated to the same cluster. **b.** Translated repeat lengths (number of amino acids) of 48 loci within CDS with at least 10 fully-translated (ORF maintained) alleles of different sizes.

Out of the 196,351 TR loci located within CDS, 16.5% (32,392) at least one allele varies in repeat number from the other alleles of the same locus. We identified 331 loci located outside of segmental duplications with at least ten different alleles by repeat length and translated their nucleotide sequences *in silico* to illustrate their extent of polymorphisms in amino acid lengths. To scrutinize if the repeat sequences were accurately assembled, we classified translations as “full” translations when the translated flanking sequences both upstream and downstream of the repeats were identical to those in the corresponding gene transcript from the GRCh38 Gencode annotations, and “partial” if otherwise. Forty-eight loci in 46 genes exhibited ten or more different fully-translated alleles within a single coding exon, and the extent of their size polymorphism in amino acid lengths are depicted in Fig. 2b. Overall, of the 24,383 alleles assembled by the various assemblers on these 48 loci in all samples, 93.2% (22,714) yielded full *in silico* translations, suggesting potentially accurate TR reconstructions in most of these assemblies. Additionally, we performed multiple sequence alignments (MSAs) for the translated alleles of each locus and selected the polymorphic CAC STR in the histidine-rich carboxyl terminus of the single-exon *HRCT1* gene to illustrate the diversity in sequence compositions in addition to the length differences that can occur in polymorphic alleles of a single locus (Supplementary Fig. 8). We note that the MSA visualization software (pyMSAviz) failed to render three loci (*HRNR*, *MUC3A*, and *MUC4*) that harbor some of the most polymorphic and longest alleles we identified. Comparing the performance of the different assemblers in combination with the two sequencing platforms, PacBio HiFi sequencing and its assemblies demonstrated a higher rate of producing full translations (97.2% for Hifiasm; 91.2% for Flye; 96.7% for Racon) than ONT sequencing and the Shasta+Hapdup assembly pipeline (86.6%) (Supplementary Fig. 9). The 48-bp VNTR in exon 2 of *MUC4* at chr3:195,778,826-195,788,662 was the most polymorphic coding TR found with over 100 alleles detected. Two other neighboring mucins on 7q22.1, *MUC3A* and *MUC12,* also harbor highly polymorphic VNTRs that occupy their largest coding exons, respectively. It is notable that eight of the highly polymorphic coding loci are associated with repeat expansion diseases: *AR*, *ATN1*, *ATXN1*, *ATXN2*, *CACNA1A*, *HTT*, *TBP*, and *THAP11* (see below).

### Motif heterogeneity

Of all STRs genotyped, 7.7% (1,029,524/13,394,896) have more than one motif sequence genotype on the same locus across samples. It has been reported that disease-associated STRs made up of non-canonical motifs are correlated with expansion sizes and pathogenicity, one of the most well-known examples being the AAAAG pentanucleotide associated with cerebellar ataxia, neuropathy and vestibular areflexia syndrome (CANVAS)^49^. Our conjecture is that such an association between motif sequences and repeat sizes may also exist in healthy populations. To expedite the search, we focused on STRs where two major motifs (that together make up at least 90% of the total reported alleles) exist that have the same length but differ in sequence composition. Testing the statistical significance of associations, we found seven loci with alleles that demonstrated such a motif-size correlation (Fig. 3, left). The allelic sequences collected for each motif were summarized in the form of sequence logos for visual confirmation of the different motif compositions (Fig. 3, right). It is interesting to note that in four cases, *TSG101*, *METAP2*, *NFS1*, *ARHGEF3*, the larger alleles are made up of long tracts of the variant motif in juxtaposition to a smaller tract composed of motifs associated with the smaller allele, suggesting insertions of repeat sequences of a slightly different motif. We also note that for *SH3RF3* and *CCDC38*, the shorter rather than the longer alleles harbor motifs different from the reference genome, unlike the previously described four cases. Although the difference between the size distributions of the two allele groups of *BEAN1*, the expansion of which is associated with spinocerebellar ataxia type 31 (SCA31), is statistically significant (p<0.05), seven of the AATAA alleles were outliers to the other shorter alleles and even longer than the rare variant CAATA alleles. All loci harboring the long *BEAN1* alleles are heterozygous, and the sizes of long alleles extracted from assemblies were confirmed by Straglr genotyping from ONT reads.

**Figure 3.**
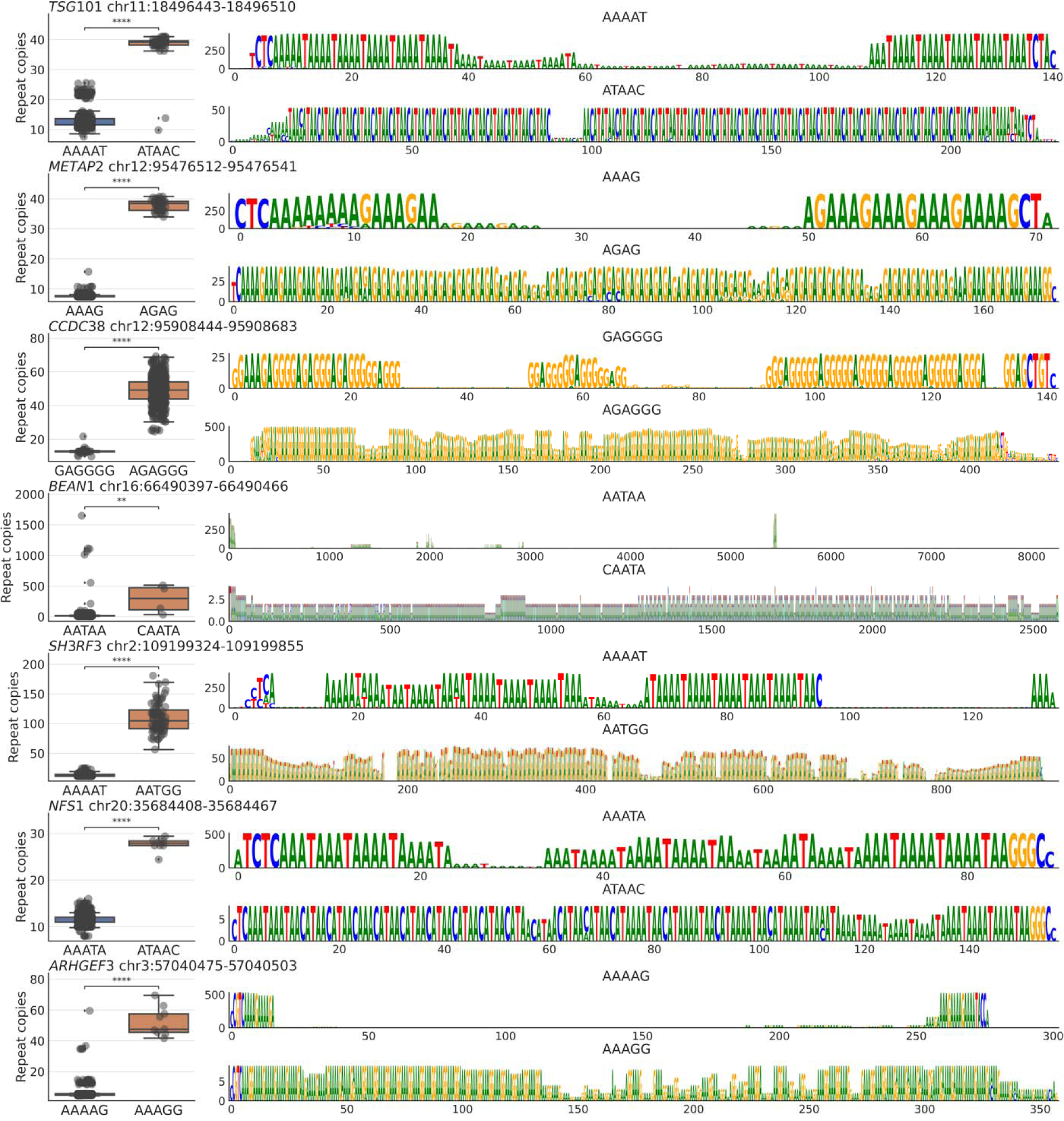
Seven genic STR loci where alleles of different motifs are associated with statistically significant differences in size distribution. (**left**) Size (repeat copies) distributions of each motif group. P-value annotation: *:0.01 < p 0.05; **: 0.001 < p 0.01; ***: 0.0001 < p 0.001; ≤ ≤ ≤ ****: p 0.0001. (**right**) Sequence logos are generated from TRs of all alleles belonging to each ≤ motif group (top and bottom panels). The Y-axis indicates the number of alleles, X-axis indicates repeat length (bp).

### Ancestry delineation

Information on the population origins of our study samples was accessible from the 1000 Genomes study, which informs that our samples originate from five superpopulations: African (AFR, n=87), European (EUR, n=24), East Asian (EAS, n=55), admixed American (AMR, n=56), and South Asian (SAS, n=50) (Supplementary table 6). While previous studies have used single nucleotide variants (SNVs) as the vehicle for genetic ancestry inferences^50,51^, we explored whether polymorphic TR genotypes alone can be used to segregate samples to their origin populations. All TRs that have all 544 haplotypes genotyped and a standard deviation of at least two repeat copies (n=782) were subjected to a principal component analysis (PCA). The mean repeat copies of the two haplotypes were used in the calculation. A clear separation can be observed between the AFR and EAS samples, while EUR and SAS samples displayed a less distinct separation, and both mingled with the admixed AMR samples (Fig. 4 left). The PCA results are similar to those performed by Frontanilla *et al*. using 22 STR genotypes from 2054 1KGP samples^52^. Interestingly, a similar but tighter clustering pattern could be observed when PCA was performed using genome-wide SNVs (n=47,426,650) called using the diploid assemblies against the GRCh38 reference (Fig. 4 right).

**Figure 4.**
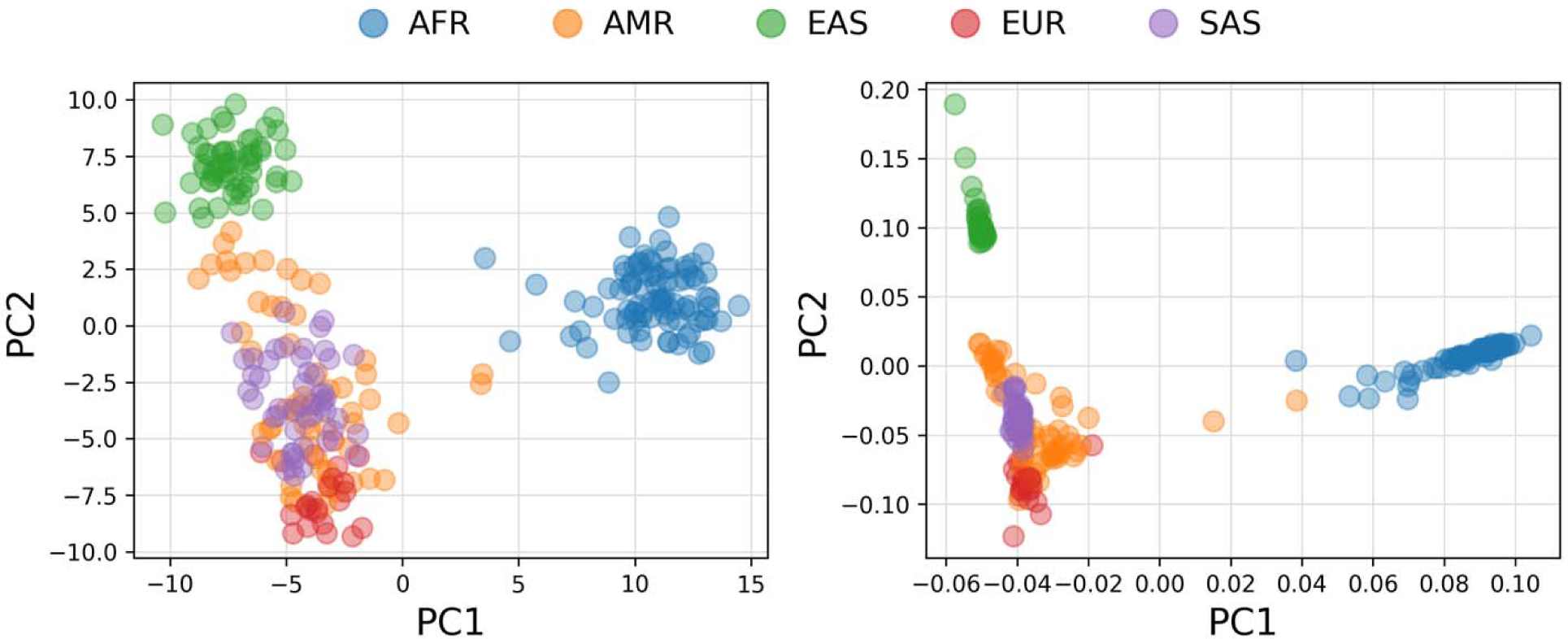
PCA of genetic ancestry using TR (left) and SNVs (right).

## Discussion

The characterization of the TR landscape in human genomes is now possible with LRS, which provides an unparalleled ability to profile TRs of all sizes and complexities. Furthermore, the high-quality diploid assemblies derived from LRS datasets facilitate robust genome-wide TR genotyping without the need to depend on pre-existing TR annotations. We have demonstrated in this study that *de novo* annotations of TRs in LRS assemblies are feasible and that the current state-of-the-art long-read assemblers produce a robust reconstruction of TR alleles, even those that are multiple kilobases long. This is evident from the high repeat size concordance between assembly-based and read-based TR genotyping results and the high degree of ORF preservations in the assembled expanded and polymorphic alleles of coding TRs after *in silico* translation.

Nonetheless, there remains room for improvement in assembly-based TR genotyping as occasional cases of potential false homozygosity were identified and supported by clear detection of alleles harbouring disparate sizes from read-alignment-based genotyping. One such example can be found at the ATTCT STR in *ATXN10*, where an expansion is associated with spinocerebellar ataxia type 10, in the HPRC sample HG01123. While the trio Hifiasm assembly reconstructed a 65 bp repeat for both the paternal and maternal alleles, Straglr reported a ∼6.2 kb allele supported by ten Nanopore and two PacBio Hifi reads in addition to the 65 bp allele (Supplementary Fig. 10). We found another potential assembly shortcoming in the occasional end of contiguity at TRs. An example is the impure CCA repeat in intron 30 of the subtelomeric gene *RTEL1*, which is successfully assembled by Hifiasm in only 60% (173/286) of all HPRC haplotypes. Complementary Straglr genotyping indicates that the missed alleles, due to truncated assemblies, were polymorphic ranging in size between 2-22 kb, suggesting that allele size may not be the determining factor for the failure of Hifiasm at this locus.

For our pipeline, we used more inclusive TRF parameters than those used for generating the UCSC [https://genome.ucsc.edu/cgi-bin/hgGateway] Simple Repeats annotation, so that many previously unannotated loci are incorporated into our catalog. Of the novel loci, many have short copy numbers (2-3) and include impure repeat units (partial or slightly variant motifs).

Nevertheless, we believe it is advantageous to include them as we found that several known disease loci were missing in the UCSC GRCh38 repeat annotations. A notable example is the *FXN* (GAA)_6_ locus (chr9:69,037,287-69,037,304), the expansion of which is the causative mutation of Friedreich ataxia. From our results in this study, we identified four loci of unannotated genic STRs with expanded alleles, which serve to disambiguate the validity of the short and impure TRs (Supplementary Fig. 11). The smallest and biggest alleles of two of them, *MORN1* AGCCC at chr1:2,335,829-2,335,846, and *SCOC* CTT at chr4:140,362,274-140,362,299, differ by more than 2kb in size. A comprehensive TR catalog will be a good companion for many current TR genotyping software programs that require the provision of coordinates and motif sequences for genome-wide detection of repeat expansions.

Long-read sequences allow us to collect many lengthy TR alleles not captured in the reference genomes. We found 87 loci, many of them in subtelomeric regions^53^, that have alleles at least 10kb larger than both the GRCh38 and T2T-CHM13 reference genomes (Supplementary Table 5). A collection of sizable TR alleles in the healthy human population is invaluable for screening for potential pathogenic expansions against normal polymorphisms that are not captured in the reference genomes. Although the number of samples used in constructing our long-read assembly-based catalog is smaller than several previous TR catalogs generated using short-read data^54,55^, the number of samples and the number of alleles collected per locus do offer significant statistical power to differentiate mutations from polymorphisms. To this end, we developed a software utility^56^ that facilitates this screening process to accompany the catalogs.

An increasing number of studies have shown that VNTRs, in addition to STRs, may have biological relevance in human phenotype expression and disease etiology. A few examples include *HRNR*^57^, *ACAN*^58^, *DRD4*^59,60^, and *TCHH*^61^ – among the most polymorphic coding VNTRs we found in this analysis. Non-coding VNTRs can also impact human health: some documented^62–64^ VNTR-disease associations include *ABCA7* and Alzheimer’s disease^65^, *WDR7* and amyotrophic lateral sclerosis^66^, and *SLC6A3* and attention deficit hyperactivity disorder^67^. The availability of genome-wide VNTR genotypes that are often absent in existing NGS-based TR catalogs will contribute to the control set used in association studies for identifying more health-related loci.

## Methods

### Catalog construction

#### Extracting assembly sequences for TR detection by TRF

TRF (v4.09) was used to extract TRs from all sequence assemblies. We found that it is infeasible to subject all individual long-read scaffold sequences for processing because TRF tends to stall in excessively repeat-laden genomic regions. To circumvent this bottleneck, we identified and screened out such problematic regions before running TRF. We postulate that these regions would correspond to assembly sequences that were unmappable or could not be mapped uniquely to the reference genome. Thus, each haplotype assembly was first mapped to GRCh38 or T2T-CHM13 by minimap2^68^ (v2.24, preset asm5). The resulting mappings, in Pairwise mApping Format (PAF), were then filtered by a minimum mapping quality of 60. The mappings were grouped by scaffold ID and sorted by alignment length, with the chromosome of the longest mapping within each group assigned as the origin chromosome. Mappings were further screened by overlapping their genome coordinates with centromere coordinates of the assigned chromosome so that scaffold regions mostly mapped (80% of length) to centromeres were skipped. Finally, mappings were overlapped against themselves to identify disparate scaffold sequences mapped to the same genomic regions (80% reciprocal intersections). Sequences corresponding to these mappings were filtered out. Retained assembly sequences were extracted using BEDTools^69^ (v2.30.0) getfasta and served as inputs for individual TRF runs.

Despite the overall effectiveness of this strategy, TRF was not able to finish processing a small number of scaffold subsequences based on the filtered mappings. For such cases, we fragmented the problematic scaffold sub-sequence into 1Mb chunks with 1kb overlaps and reran to completion.

#### Post-processing of TRF results

TRF was run with the parameters: “2 5 5 80 10 10 500 -d -h”. Coordinates of the TRs found in the extracted sub-sequences described above were converted back to the original scaffold coordinates for downstream processing. Homopolymers and TRs with motifs longer than 100bp were discarded. Because TRF results contain a lot of overlapping calls, we screened the results to reduce the redundancy of TRs in the final generated catalog. TRF calls were first grouped by scaffolds and then sorted by their start and end coordinates. While iterating through the calls, overlapping TRs were identified when one of the following three conditions was met: (1) The coordinates of the current call are subsumed in the coordinates of the previous call or vice versa; (2) The overlapping size is at least 80% of the spans of the current and previous calls, or (3) The distance between the start and end coordinates of two calls is less than 5 bp. The TR call of the overlapping pair with a better alignment score (field 8 of the TRF .dat output), or a shorter motif (field 2 of the TRF .dat output) for tiebreaking, was kept and used for comparison against the next TR call. The final list of retained TRF calls was translated to TSV format for the next step of mapping to the reference genome.

#### Mapping TRs from assembly to the reference genome

To map TRs detected from the assembly onto the reference genome, 500 bp flanking sequences on either side of each TR were extracted from the assembly and aligned against the reference genome using BWA mem^70^ (v0.7.12). TRs located < 500 bp to the ends of the scaffold were skipped. TRs were considered successfully mapped when the flanking sequence alignments passed all of the following conditions: (1) Both flanks were mapped to the same chromosome; (2) The orientation of one flank was opposite to the other, i.e., they point towards each other; (3) No more than 10 bp of the outer edges (ends farthest to the repeat) of the flanking sequences were unmapped (alignments of the inner edges flanking the repeat were not checked due to possible errors in ascertaining repeat boundaries); (4) The deduced end reference genome coordinate was larger than the start coordinate; (5) In order to reduce egregious mapping errors, the reference genome span determined from the flanking sequence alignments was no more than ten times the repeat size detected in the assembly. The genomic locations together with their corresponding assembly positions of each successfully mapped TR were tracked and were subject to a final quality check against the minimap2 mapping results of the assemblies. The genome location of each TR was checked to ensure that it resided within the mapped genomic span of its origin scaffold sequence; if not, the TR was excluded from the final list. The origin of each genotype (sample name and haplotype) was affixed to each TR in the final BED output to facilitate the final integration step. Parental haplotypes were designated “paternal” or “maternal” for the HPRC trio Hifisam assemblies; “hap1/hap2”, “h1/h2”, and “p1/p2” for the HPRC default Hifiasm assemblies generated in-house, HGSVC2, and 1KGP ONT assemblies, respectively for which the parental origins of the assembled sequences were not ascertained. Different initials were used to differentiate the sources of the sequence data.

#### Editing and supplementing assembly-based TR genotypes with Straglr results

ONT and HiFi sequencing reads were first aligned against the reference genome (GRCh38 or T2T-CHM13) with minimap2. The resulting alignments were then processed by Straglr (v1.4.1) to capture TRs with 2-100 bp motif lengths and sized at least 100 bp larger than the reference genome with at least two supporting reads. Straglr results were then intersected using BEDTools with the genotyping results from both haploid assemblies to associate identical loci detected from both analyses. Loci on the same chromosome with at least 80% reciprocal overlaps in their coordinate spans and uniquely mapped between Straglr and each haplotype were retained for further comparison. The calls from the two methods were considered to agree when the sizes for a given allele were within 10% of each other. For loci with disagreeing results or loci with a missing allele from either haploid assembly, Straglr results were used to replace the assembly-based call (edits) or fill in the missing genotype (supplements) under one of the following conditions: (a) When both ONT and HiFi sequencing reads were available, allele(s) called by Straglr on data from both technologies agreed with each other; or (b) When Straglr results were only available from one technology (either because only one long-read data type is available for a particular sample or the locus was not identified by Straglr to have an expansion in one data type), the allele called by Straglr was adopted if there were at least ten support reads when the allele size was <= 5kb or four supporting reads when the allele size was > 5kb. The different requirements for supporting evidence were enforced because larger alleles were usually captured in their entirety by fewer sequencing reads. Expanded loci reported by Straglr’s genome scan using data from either technology not captured by the assembly were included when individual alleles had the minimum read support level, as described above. For cases of either edit or supplement, genotypes from HiFi data were chosen over ONT genotypes if Straglr results agreed with each other because repeat sizes from individual HiFi support reads tend to exhibit more homogeneous repeat size distributions.

#### Combining data from individual haplotypes of all samples

The list of TRs from every sample haplotype was overlapped with all of the others using BEDTools intersect based on their reference genome coordinates, requiring a reciprocal overlap (-r) of at least 70% (-f 0.7). To expedite this multitude of comparisons, we split each list of TRs by their cytoband locations for parallel processing. All identified pairwise overlaps were then used to create edges in an undirected graph with the assumption that each cluster of connected components represented genotypes originating from the same locus. For each genotype group, the most frequently observed coordinates and motif sequence were picked as the consensus position and motif to represent the underlying TR locus. Two motif sequences were considered the same if any permutation of one was identical to the other. In situations where there was more than one genotype call from the same haplotype present in the cluster, the call that had identical coordinates with the consensus position or the one that had the highest overlap with the consensus position was taken. The copy numbers in each haplotype in the final list, reported in one decimal float number, were re-calculated by dividing the repeat size by the length of the consensus motif. In addition to the sizes and copy numbers, frequencies of distinct motifs observed and the size difference of the largest allele from the reference were reported for each locus.

### Reference sequence and annotation data

The GRCh38 genome FASTA was generated by concatenating the chromosome sequences (chr1-22, chrX and chrY) downloaded from the UCSC Genome Browser: http://hgdownload.soe.ucsc.edu/goldenPath/GRCh38/chromosomes/. Gencode (release 43) was used as the gene annotation for GRCh38: https://ftp.ebi.ac.uk/pub/databases/gencode/Gencode_human/release_43/gencode.v43.annotation. gtf.gz. For the T2T-CHM13 reference, the genome FASTA (v2.0), chain files for liftOver genome coordinates (grch38-chm13v2.chain and chm13v2-grch38.chain), and gene annotations (NCBI RefSeqv110) were downloaded from the Telomere-to-telomere consortium CHM13 project (https://github.com/marbl/chm13). Both GTFs were sorted by BEDTools, compressed by BGZIP (v1.10.2), and indexed by Tabix^71^ (v1.10.2). Other annotations such as Simple repeats, RepeatMasker, Centromeres, Chromosome bands, and Gaps were downloaded from the “Repeats” and “Mapping and Sequencing” groups of both GRCh38 and “T2T CHM13v2.0” assemblies using the Table Browser utility of the UCSC Genome Browser.

### LiftOver between GRCh38 and T2T-CHM13 coordinates

LiftOver was performed using the command: liftOver -bedPlus=5 source.bed source-target.chain target.bed target.failed.bed, where source and target are GRCh38 and chm13 coordinates, respectively, or vice versa.

### Association of TRs between GRCh38 and T2T-CHM13 catalogs

Coordinates (chromosome, start, end) of all loci with from either the GRCh38 or T2T-CHM13 catalog were saved in BED format, with an additional field added to preserve the source coordinate in a UCSC-format label - “chromosome:start-end”. This source BED was lifted over to the target genome as described above, and a two-column file composed of the target and source UCSC-format genome coordinates of the loci with successful conversion was generated. We then used the UNIX join command to link the first field of the GRCh38-to-T2T-CHM13 file (i.e., liftOver coordinates from the GRCh38 catalog) to the second field of the T2T-CHM13-to-GRCh38 files (i.e., source coordinates from the T2T-CHM13 catalog) to identify all the equivalent loci.

### Identification of TRs with potential size error in one reference and not the other

A BED file composed of all loci from the catalog was generated as the source file. This file was lifted over to the target genome coordinates as described above. The start and end coordinates were used to calculate the TR sizes in the source and target genomes, respectively. The mean and SD were calculated for all allele sizes genotyped for each locus. A maximum SD (SD_max_) was set as 10 bp and all loci with SD > SD_max_ were removed. Size differences between the mean size of each locus and the TR size in both source and target genomes were calculated. Loci with an absolute size difference <= SD_max_ for the source and not the target genome were ones where the sizing in the source is potentially more accurate than the target reference genome. This analysis was repeated using either the GRCh38 or the T2T-CHM13 catalog as the source coordinates.

### FASTQ generation

BAM files downloaded from HPRC and HGSVC2 were converted to FASTQs using the bamtofastq utility from BEDTools: bedtools bamtofastq -i <BAM> -fq /dev/stdout | gzip > <FASTQ.gz>, where <BAM> was the input BAM, and <FASTQ.gz> was the compressed FASTQ after conversion. Alignment BAMs from 1KPG-ONT were available and directly used as input for Straglr processing.

### Aligning sequencing reads to the reference genome

FASTQs of Nanopore or PacBio (HiFi or CLR) sequencing reads for each sample were aligned to the GRCh38 and T2T-CHM13 reference genome by minimap2 and converted to BAM using the following command: minimap2 -ax <PRESET> -t 16 -Y -L –MD <genome.fa> <FASTQS= | samtools view -bhS - | samtools sort -m75G - -o <BAM>, where <PRESET> = map-ont or map-pb for Nanopore or PacBio reads, respectively, <FASTQS= was the space-limited list of the full-paths of the input FASTQs, and <BAM> was the output BAM.

### Mapping assembly to the reference genome

Assembled FASTAs were mapped to the reference genome by minimap2 using the following command: minimap2 <genome.fa> <asm.fa> -x asm5 -t 42 > <PAF>, where <genome.fa> and <asm.fa> were the reference genome and haplotype FASTAs, respectively, and <PAF> was the output PAF.

### Hifiasm assembly

FASTQs of HiFi sequencing reads were assembled by Hifiasm (v0.19.5) in default mode using the following command: hifiasm -o <SAMPLE_NAME> -t 32 <FASTQS=, where <SAMPLE_NAME> was the prefix of the output files and <FASTQS= was the space-limited list of the full-paths of the input FASTQs. Each of the two assembled haplotype GFAs was converted into FASTA by the following awk command: awk ’/^S/{print “>”$2;print $3}’ <GFA> > <FASTA>. All assembly FASTAs (including the downloaded ones from HPRC) were compressed by BGZIP and indexed by Samtools^72^ (samtools faidx).

### Straglr runs

Straglr (v1.3.1) was run in genome-scan mode on alignment BAMs with the following parameters: straglr.py

<BAM> <genome.fa> <PREFIX> --min_str_len 2 --max_str_len 100 -- min_ins_size 100 --genotype_in_size --min_support 2 --min_cluster_size 2 --max_num_clusters 2 -- exclude <exclude.bed> --nprocs 32, where <BAM> was the alignment bam, <genome.fa> was the reference genome fasta, and <exclude.bed> contains coordinates of the following genomic regions that were skipped for scanning: 1) simple tandem repeats duplications; 3) centromeres; and 4) sequencing gaps. ^≥^ 10kb; 2) segmental

### Comparison of genotyping results from Nanopore, HiFi, and Hifiasm

Samples with Straglr results from both Nanopore and HiFi sequencing data and genotype results from Hifiasm were analyzed. We first used BEDTools to overlap (bedtools intersect -f 0.8 -r) the TR coordinates from each sample’s Straglr and Hifiasm-based results to identify matching loci. Of the matching loci, we compared the repeat size of each Straglr allele against Hifiasm alleles. Straglr alleles with too little read support (< 4 or <10% of read support of the other allele) were not used for comparison. As Straglr only reports one allele for homozygous loci, the same allele was used twice for comparison against Hifiasm haplotypes in such cases. Alleles with a size difference <= 10% between Straglr and Hifiasm alleles were considered to be matching. A Boolean matching result of each Hifiasm allele against both Nanopore and HiFi Straglr results was tallied in addition to the allele size. The results of all samples were pooled for visualization using the Python UpSetPlot^73^ package (https://github.com/jnothman/UpSetPlot).

### Clustering of alleles based on repeat copies

Repeat copies of all alleles genotyped for a given TR were first sorted numerically in ascending order. The first (i.e., smallest) value was taken as the first member of a cluster. Starting from the second value until the last, each value was added to the last cluster if the value was bigger than the previous one by less than 5% of the smaller value. Two values that differed by less than one repeat copy were automatically placed into the same cluster. A new cluster was started with the current value if the conditions above were not fulfilled. All clusters with fewer than 10 values were discarded to reject outliers.

### Inspection of polymorphic coding TRs

Assembled sequences of selected TRs embedded within single exons of CDS were extracted from assembly scaffolds using the Python Pysam module (v0.19.1). All extracted sequences were first standardized to correspond to the positive strand of the reference GRCh38 genome. They were then reverse-complemented if the mapped gene was located on the negative strand of the reference genome. To determine the frame for translating the nucleotide sequence, we first identified transcripts that fulfilled the following conditions: 1) The start and end genomic coordinates of the repeat were embedded within a single exon; 2) The transcript had both start and stop codons. The most common start codon of all transcripts that met the above conditions was used to determine the frame for translating the nucleotide sequence. The transcript distance of the repeat from the start codon was calculated by summing the distance between the start codon and its downstream exon boundary, the sum of all exon lengths between the start codon and the repeat, and the distance of the repeat start to its upstream exon boundary. The remainder after dividing the transcript distance from the start codon by three was used to derive the frame for translation. The sequences were then translated to amino acids using the BioPython (v1.79, https://biopython.org/) package. Multiple sequence alignments of the translated repeat sequences were performed using Clustal Omega^74^ (v1.2.4): clustalo --wrap=150000 --output-order=input-order -i <in.fa> -t Protein -o <out.fa>, where <in.fa> was the FASTA of pooled repeat amino acid sequences for each locus, and <out.fa> was the FASTA of aligned sequences. The alignments were converted to an image for visualization using the PyMSAviz package (https://github.com/moshi4/pyMSAviz).

### Motif sequence and size correlation

TRs with motifs 2-6 bp in length were screened to contain multiple motifs of the same length that made up ≥ 90% of all alleles genotyped. Alleles were segregated by their motifs, and each pairwise comb^≥^ination of allele groups was tested for statistical significance (P-value < 0.05) in their size differences measured in repeat copies using the two-sided Mann-Whitney U test, implemented by the SciPy Statistics package. Because we employed slightly error-tolerant TRF parameters in capturing TRs to construct the catalog, we re-ran TRF on the haplotype sequences of every allele of all the top-ranked (by P-value) cases using more stringent parameters (2 7 7 80 10 50 500) to make sure the reported catalog motif remained the predominant motif with increased stringency. The size distribution differences between motifs of verified loci were annotated and visualized using the Python statannot^75^ package. For each reported locus, the TR sequences of all alleles were extracted from their respective assemblies with 5-bp flanking sequences affixed, reverse-complemented if the scaffold sequence was previously determined to originate from the negative strand of the reference genome, pooled for multiple sequence alignment using Clustal Omega (v1.2.4): clustalo --wrap=150000 --output-order=input-order -i <in.fa> -t DNA -o <out.fa>, where <in.fa> was the FASTA of pooled motif sequences, and <out.fa> was the FASTA of aligned sequences. The output sequences were converted to sequence logos using the Python Logomaker package^76^ for visual comparison of sequences between motif groups of the reported loci.

### PCA using repeat sizes

Loci with both haplotypes genotyped in all samples and a standard deviation of at least two repeat copies was identified. The mean repeat copy of the two haplotypes was used as the representative genotype of a single locus for a given sample. A 2-dimensional matrix was compiled such that each row is a vector containing the genotypes of all identified loci for an individual locus. The matrix was normalized using the StandardScaler utility of the preprocessing module and then fed for fitting and dimension reduction using the PCA module, both from the Python scikit-learn package (v1.3.1).

### PCA using single nucleotide variants

Each assembled haplotype was mapped to GRCh38 using minimap2 and variations (single nucleotide changes, insertions, and deletions) were identified in the resultant PAF output using the paftools.js script: minimap2 -cx asm5 --cs <genome.fa> <haplotype.fa> | sort -k6,6 -k8,8n | paftools.js call -f <genome.fa> - > haplotype.vcf. The variant calls from the two haplotypes of each sample were merged using bcftools merge, and the genotype of each variant in the merged VCF were transformed into “0|1” or “1|0” if one of the two haplotypes harbored the variant and into “1|1” if both haplotypes harbored the variant, creating phased VCFs for each sample. The phased VCFs of all the samples were then merged using bcftools merge, and bi-allelic single nucleotide variants in chr 1-22 and chrX were extracted from the merged VCF using bcftools view. PCA was then performed on these variants using plink (v2.00-10252019-avx2)^77^.

## Data availability

BED files of TR genotypes with sample origins, sizes, repeat copies, and annotated features for both GRCh38 and T2T-CHM13 genome references together with software developed for generating the catalogs are available from the GitHub repository: https://github.com/bcgsc/tr_catalog. Online locations of LRS read and assembly sequences used in this study can be found in Supplementary Table 1.

## Supporting information

Supplementary inoformation

## Data Availability

All data produced are available online at https://zenodo.org/records/11522276

https://s3-us-west-2.amazonaws.com/human-pangenomics/index.html?prefix=working/

https://ftp.1000genomes.ebi.ac.uk/vol1/ftp/data_collections/HGSVC2/release/v1.0/

https://s3.amazonaws.com/1000g-ont/index.html?prefix=FIRST_100_FREEZE/

## Acknowledgements

This study was supported by the Canadian Institutes of Health Research (CIHR) Project Grant (PJT-169074). I.-S.R.-B. is funded by the CIHR Research Excellence, Diversity, and Independence (REDI) Early Career Transition Award (DI2-190730). The authors thank the Digital Research Alliance of Canada for the Resource Allocation that supported some of the analyses performed in this study.

## References

1. Ziaei Jam, H., et al. A deep population reference panel of tandem repeat variation. Nat. Commun. 14, 6711 (2023).

2. Course, M. M., Sulovari, A., Gudsnuk, K., Eichler, E. E. & Valdmanis, P. N. Characterizing nucleotide variation and expansion dynamics in human-specific variable number tandem repeats. Genome Res. 31, 1313–1324 (2021).

3. Gall-Duncan, T., Sato, N., Yuen, R. K. C. & Pearson, C. E. Advancing genomic technologies and clinical awareness accelerates discovery of disease-associated tandem repeat sequences. Genome Res. 32, 1–27 (2022).

4. English, A. C. et al. Analysis and benchmarking of small and large genomic variants across tandem repeats. Nat. Biotechnol. 1–12 (2024) doi:10.1038/s41587-024-02225-z.

5. Lu, T.-Y., Human Genome Structural Variation Consortium & Chaisson, M. J. P. Profiling variable-number tandem repeat variation across populations using repeat-pangenome graphs. Nat. Commun. 12, 4250 (2021).

6. Rajan-Babu, I.-S., Dolzhenko, E., Eberle, M. A. & Friedman, J. M. Sequence composition changes in short tandem repeats: heterogeneity, detection, mechanisms and clinical implications. Nat. Rev. Genet. 1–24 (2024) doi:10.1038/s41576-024-00696-z.

7. Homepage. https://harrietdashnow.com/STRchive/.

8. Halman, A., Dolzhenko, E. & Oshlack, A. STRipy: A graphical application for enhanced genotyping of pathogenic short tandem repeats in sequencing data. Hum. Mutat. 43, 859–868 (2022).

9. Pathogenic Short Tandem Repeats | gnomAD v3.1.2 | gnomAD. https://gnomad.broadinstitute.org/short-tandem-repeats?dataset=gnomad_r3.

10. Trost, B. et al. Genome-wide detection of tandem DNA repeats that are expanded in autism. Nature 586, 80–86 (2020).

11. Mojarad, B. A. et al. Genome-wide tandem repeat expansions contribute to schizophrenia risk. Mol. Psychiatry 27, 3692–3698 (2022).

12. Erwin, G. S. et al. Recurrent repeat expansions in human cancer genomes. Nature 613, 96–102 (2023).

13. Mitina, A. et al. Genome-wide enhancer-associated tandem repeats are expanded in cardiomyopathy. EBioMedicine 101, 105027 (2024).

14. Birnbaum, R. Rediscovering tandem repeat variation in schizophrenia: challenges and opportunities. Transl. Psychiatry 13, 402 (2023).

15. Jakubosky, D. et al. Properties of structural variants and short tandem repeats associated with gene expression and complex traits. Nat. Commun. 11, 2927 (2020).

16. Margoliash, J. et al. Polymorphic short tandem repeats make widespread contributions to blood and serum traits. Cell Genomics 3, 100458 (2023).

17. Bakhtiari, M., Shleizer-Burko, S., Gymrek, M., Bansal, V. & Bafna, V. Targeted genotyping of variable number tandem repeats with adVNTR. Genome Res. 28, 1709–1719 (2018).

18. Willems, T. et al. Genome-wide profiling of heritable and de novo STR variations. Nat. Methods 14, 590–592 (2017).

19. Fotsing, S. F. et al. The impact of short tandem repeat variation on gene expression. Nat. Genet. 51, 1652–1659 (2019).

20. Horton, C. A. et al. Short tandem repeats bind transcription factors to tune eukaryotic gene expression. Science 381, eadd1250 (2023).

21. Wright, S. E. & Todd, P. K. Native functions of short tandem repeats. eLife 12, e84043.

22. Tanudisastro, H. A., Deveson, I. W., Dashnow, H. & MacArthur, D. G. Sequencing and characterizing short tandem repeats in the human genome. Nat. Rev. Genet. (2024) doi:10.1038/s41576-024-00692-3.

23. Coster, W. D. et al. Medically relevant tandem repeats in nanopore sequencing of control cohorts. 2024.03.06.24303700 Preprint at 10.1101/2024.03.06.24303700 (2024).

24. Dolzhenko, E. et al. ExpansionHunter Denovo: a computational method for locating known and novel repeat expansions in short-read sequencing data. Genome Biol. 21, 102 (2020).

25. Dolzhenko, E. et al. ExpansionHunter: a sequence-graph-based tool to analyze variation in short tandem repeat regions. Bioinformatics 35, 4754–4756 (2019).

26. Dashnow, H. et al. STRetch: detecting and discovering pathogenic short tandem repeat expansions. Genome Biol. 19, 121 (2018).

27. Dashnow, H. et al. STRling: a k-mer counting approach that detects short tandem repeat expansions at known and novel loci. Genome Biol. 23, 257 (2022).

28. Mousavi, N., Shleizer-Burko, S., Yanicky, R. & Gymrek, M. Profiling the genome-wide landscape of tandem repeat expansions. Nucleic Acids Res. 47, e90 (2019).

29. Tankard, R. M. et al. Detecting Expansions of Tandem Repeats in Cohorts Sequenced with Short-Read Sequencing Data. Am. J. Hum. Genet. 103, 858–873 (2018).

30. Rajan-Babu, I.-S. et al. Genome-wide sequencing as a first-tier screening test for short tandem repeat expansions. Genome Med. 13, 126 (2021).

31. Depienne, C. & Mandel, J.-L. 30 years of repeat expansion disorders: What have we learned and what are the remaining challenges? Am. J. Hum. Genet. 108, 764–785 (2021).

32. Dolzhenko, E. et al. Characterization and visualization of tandem repeats at genome scale. Nat. Biotechnol. 1–9 (2024) doi:10.1038/s41587-023-02057-3.

33. Chen, S. et al. A genomic mutational constraint map using variation in 76,156 human genomes. Nature 625, 92–100 (2024).

34. MacDonald, J. R., Ziman, R., Yuen, R. K. C., Feuk, L. & Scherer, S. W. The Database of Genomic Variants: a curated collection of structural variation in the human genome. Nucleic Acids Res. 42, D986–992 (2014).

35. Nurk, S. et al. The complete sequence of a human genome. Science 376, 44–53 (2022).

36. Liao, W.-W. et al. A draft human pangenome reference. Nature 617, 312–324 (2023).

37. Aganezov, S. et al. A complete reference genome improves analysis of human genetic variation. Science 376, eabl3533 (2022).

38. Ebert, P. et al. Haplotype-resolved diverse human genomes and integrated analysis of structural variation. Science 372, eabf7117 (2021).

39. Gustafson, J. A. et al. Nanopore sequencing of 1000 Genomes Project samples to build a comprehensive catalog of human genetic variation. 2024.03.05.24303792 Preprint at 10.1101/2024.03.05.24303792 (2024).

40. Cheng, H., Concepcion, G. T., Feng, X., Zhang, H. & Li, H. Haplotype-resolved de novo assembly using phased assembly graphs with hifiasm. Nat. Methods 18, 170–175 (2021).

41. Kolmogorov, M., Yuan, J., Lin, Y. & Pevzner, P. A. Assembly of long, error-prone reads using repeat graphs. Nat. Biotechnol. 37, 540–546 (2019).

42. Vaser, R., Sović, I., Nagarajan, N. & Šikić, M. Fast and accurate de novo genome assembly from long uncorrected reads. Genome Res. 27, 737–746 (2017).

43. Kolmogorov, M. et al. Scalable Nanopore sequencing of human genomes provides a comprehensive view of haplotype-resolved variation and methylation. 2023.01.12.523790 Preprint at 10.1101/2023.01.12.523790 (2023).

44. Nurk, S. et al. The complete sequence of a human genome. Science 376, 44–53 (2022).

45. Benson, G. Tandem repeats finder: a program to analyze DNA sequences. Nucleic Acids Res. 27, 573–580 (1999).

46. Chiu, R., Rajan-Babu, I.-S., Friedman, J. M. & Birol, I. Straglr: discovering and genotyping tandem repeat expansions using whole genome long-read sequences. Genome Biol. 22, 224 (2021).

47. Hinrichs, A. S. et al. The UCSC Genome Browser Database: update 2006. Nucleic Acids Res. 34, D590–D598 (2006).

48. Cheng, H. et al. Haplotype-resolved assembly of diploid genomes without parental data. Nat. Biotechnol. 40, 1332–1335 (2022).

49. Cortese, A. et al. Biallelic expansion of an intronic repeat in RFC1 is a common cause of late-onset ataxia. Nat. Genet. 51, 649–658 (2019).

50. Bollas, A. E. et al. SNVstory: inferring genetic ancestry from genome sequencing data. BMC Bioinformatics 25, 76 (2024).

51. Warren, R. L., Coombe, L., Wong, J., Kazemi, P. & Birol, I. Human ancestry inference at scale, from genomic data. 2024.03.26.586646 Preprint at 10.1101/2024.03.26.586646 (2024).

52. Frontanilla, T. S., Valle-Silva, G., Ayala, J. & Mendes-Junior, C. T. Open-Access Worldwide Population STR Database Constructed Using High-Coverage Massively Parallel Sequencing Data Obtained from the 1000 Genomes Project. Genes 13, 2205 (2022).

53. Linthorst, J. et al. Extreme enrichment of VNTR-associated polymorphicity in human subtelomeres: genes with most VNTRs are predominantly expressed in the brain. Transl. Psychiatry 10, 369 (2020).

54. Ziaei Jam, H., et al. A deep population reference panel of tandem repeat variation. Nat. Commun. 14, 6711 (2023).

55. Cui, Y. et al. A genome-wide spectrum of tandem repeat expansions in 338,963 humans. Cell 187, 2336–2341.e5 (2024).

56. straglr/straglr_compare.py at master · bcgsc/straglr. https://github.com/bcgsc/straglr/blob/master/straglr_compare.py.

57. Lu, T.-Y., Smaruj, P. N., Fudenberg, G., Mancuso, N. & Chaisson, M. J. P. The motif composition of variable number tandem repeats impacts gene expression. Genome Res. 33, 511–524 (2023).

58. Öz, T. et al. ACAN Gene VNTR Polymorphism and Intervertebral Disc Degeneration in a Turkish Population. Med. Bull. Haseki 58, 309–314 (2020).

59. Paquet, C. et al. Dopamine D4 receptor gene polymorphism (DRD4 VNTR) moderates real-world behavioural response to the food retail environment in children. BMC Public Health 21, 145 (2021).

60. Uysal, M. A., Sever, Ü., Nursal, A. F. & Pehl van, S. Dopamine D4 Receptor Gene Exonİ III VNTR Variant Influences Smoking Status in Turkish Population. *Arch*. Neuropsychiatry 56, 248–252 (2019).

61. Mukamel, R. E. et al. Protein-coding repeat polymorphisms strongly shape diverse human phenotypes. 2021.01.19.427332 Preprint at 10.1101/2021.01.19.427332 (2021).

62. Marshall, J. N. et al. Variable number tandem repeats – Their emerging role in sickness and health. Exp. Biol. Med. 246, 1368–1376 (2021).

63. Vogan, K. VNTRs and disease risk. Nat. Genet. 55, 1421–1421 (2023).

64. Brookes, K. J. The VNTR in complex disorders: The forgotten polymorphisms? A functional way forward? Genomics 101, 273–281 (2013).

65. De Roeck, A. et al. An intronic VNTR affects splicing of ABCA7 and increases risk of Alzheimer’s disease. Acta Neuropathol. (Berl.) 135, 827–837 (2018).

66. Course, M. M. et al. Evolution of a Human-Specific Tandem Repeat Associated with ALS. Am. J. Hum. Genet. 107, 445–460 (2020).

67. Kuc, K., Bielecki, M., Racicka-Pawlukiewicz, E., Czerwinski, M. B. & Cybulska-Klosowicz, A. The SLC6A3 gene polymorphism is related to the development of attentional functions but not to ADHD. Sci. Rep. 10, 6176 (2020).

68. Li, H. New strategies to improve minimap2 alignment accuracy. Bioinformatics 37, 4572–4574 (2021).

69. Quinlan, A. R. & Hall, I. M. BEDTools: a flexible suite of utilities for comparing genomic features. Bioinformatics 26, 841–842 (2010).

70. Li, H. Aligning sequence reads, clone sequences and assembly contigs with BWA-MEM. Preprint at 10.48550/arXiv.1303.3997 (2013).

71. Li, H. Tabix: fast retrieval of sequence features from generic TAB-delimited files. Bioinformatics 27, 718–719 (2011).

72. Danecek, P. et al. Twelve years of SAMtools and BCFtools. GigaScience 10, giab008 (2021).

73. Lex, A., Gehlenborg, N., Strobelt, H., Vuillemot, R. & Pfister, H. UpSet: Visualization of Intersecting Sets. IEEE Trans. Vis. Comput. Graph. 20, 1983–1992 (2014).

74. Sievers, F. & Higgins, D. G. Clustal Omega for making accurate alignments of many protein sequences. Protein Sci. 27, 135–145 (2018).

75. Charlier, F., et al. trevismd/statannotations: v0.5. Zenodo 10.5281/ZENODO.7213391 (2022).

76. Tareen, A. & Kinney, J. B. Logomaker: beautiful sequence logos in Python. Bioinformatics 36, 2272–2274 (2020).

77. Purcell, S. et al. PLINK: A Tool Set for Whole-Genome Association and Population-Based Linkage Analyses. Am. J. Hum. Genet. 81, 559–575 (2007).

