## Supplementary inoformation for "A comprehensive tandem repeat catalog of the human genome"

Supplementary Information

### Supplementary Figures

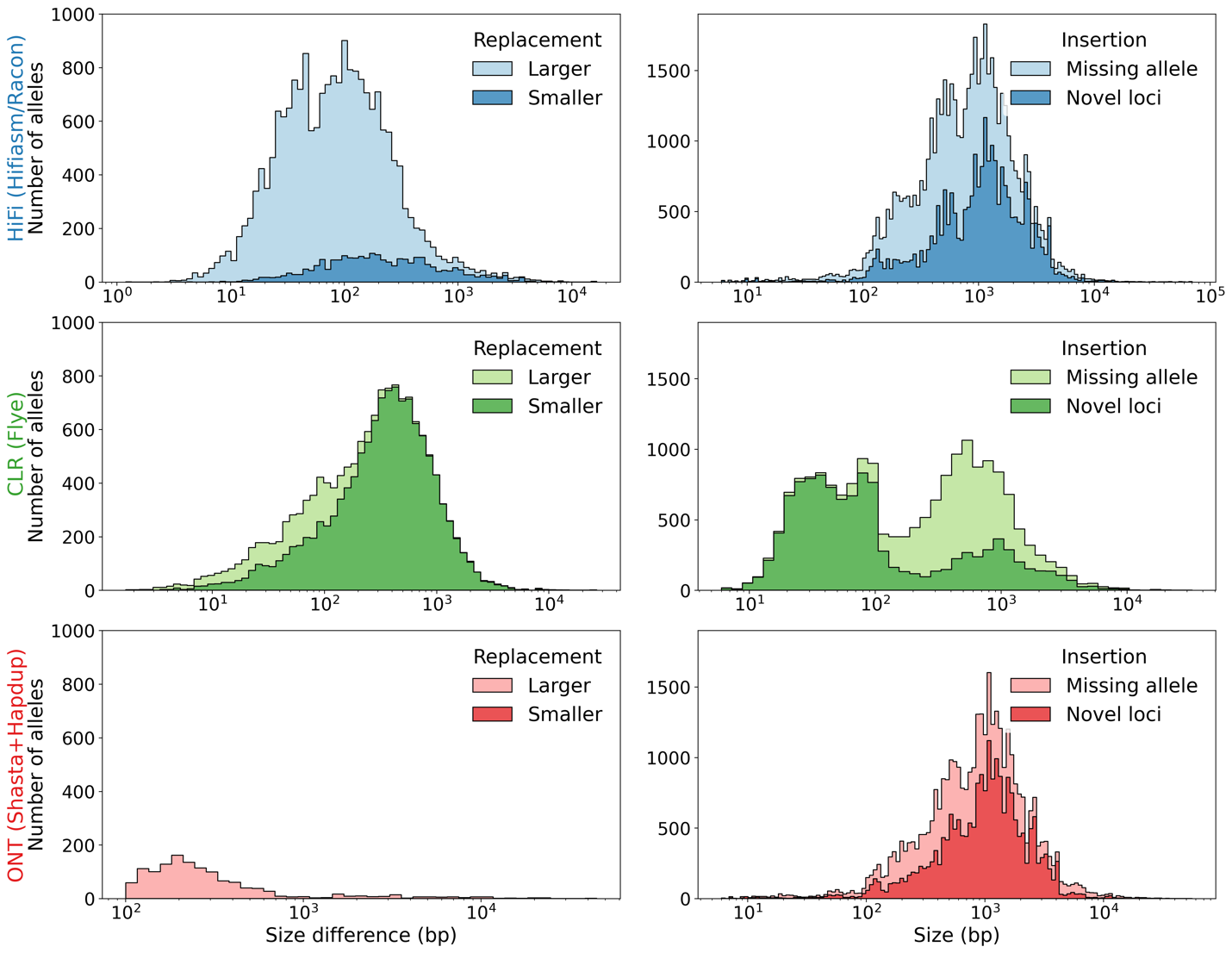

Supplementary Figure 1. Size distributions of edited alleles performed on assembly-based genotypes. Each row shows edits performed on data coming from different sequencing platforms with assembler software indicated (blue = PacBio HiFi; green = PacBio CLR; red = ONT). (Left) Alleles differentiated by whether replacements (Straglr-based) are larger (light-color) or smaller (dark-color) than original (assembly-based) genotypes. (Right) Inserted alleles (Straglr-based genotypes) composed of missing assembly haplotypes (light-color) or loci (dark-color).

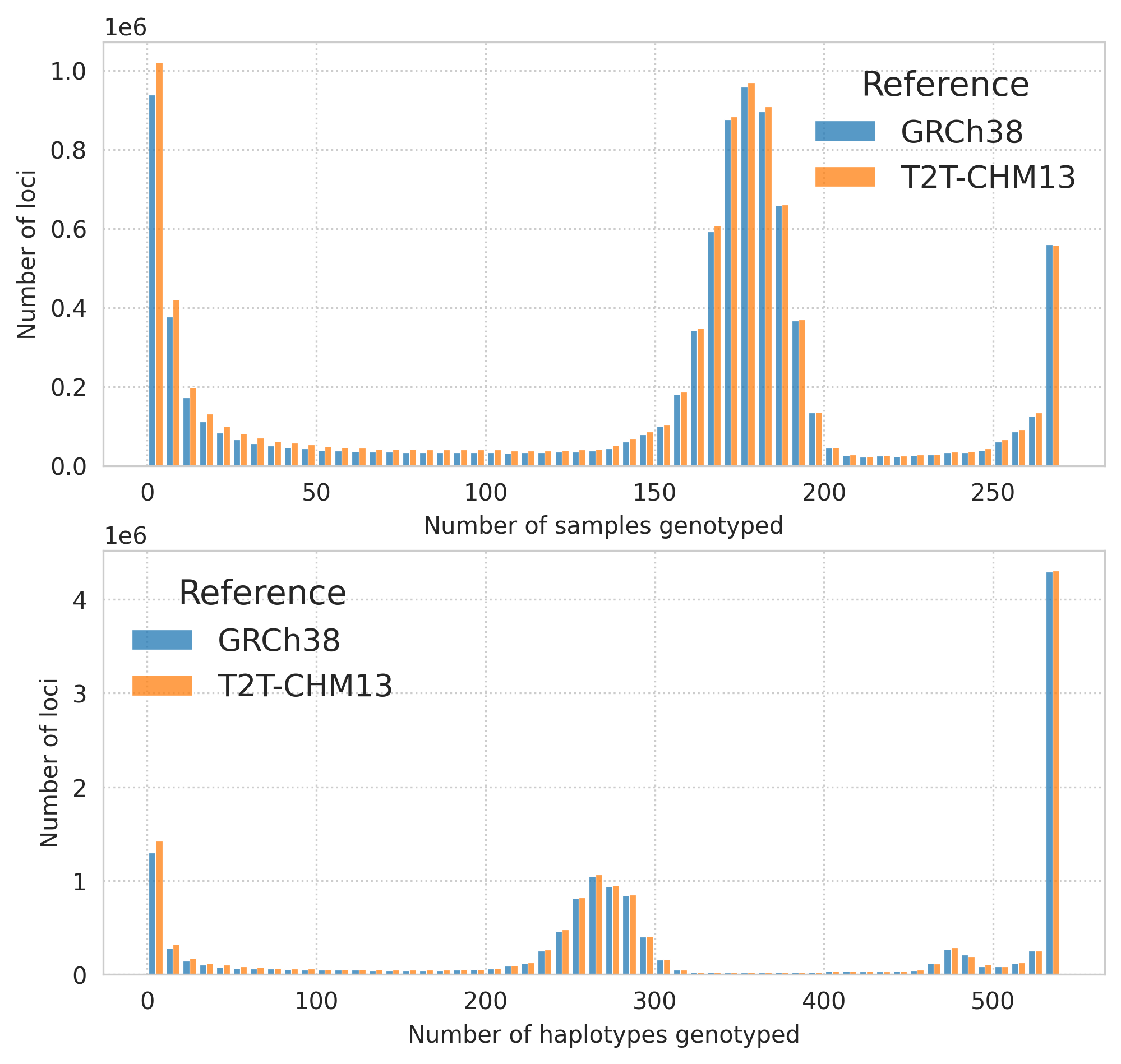

Supplementary Figure 2. Number of genotypes collected per locus. Number of loci (Y-axis) with genotypes from X(-axis) number of samples (top) or haplotypes (bottom).

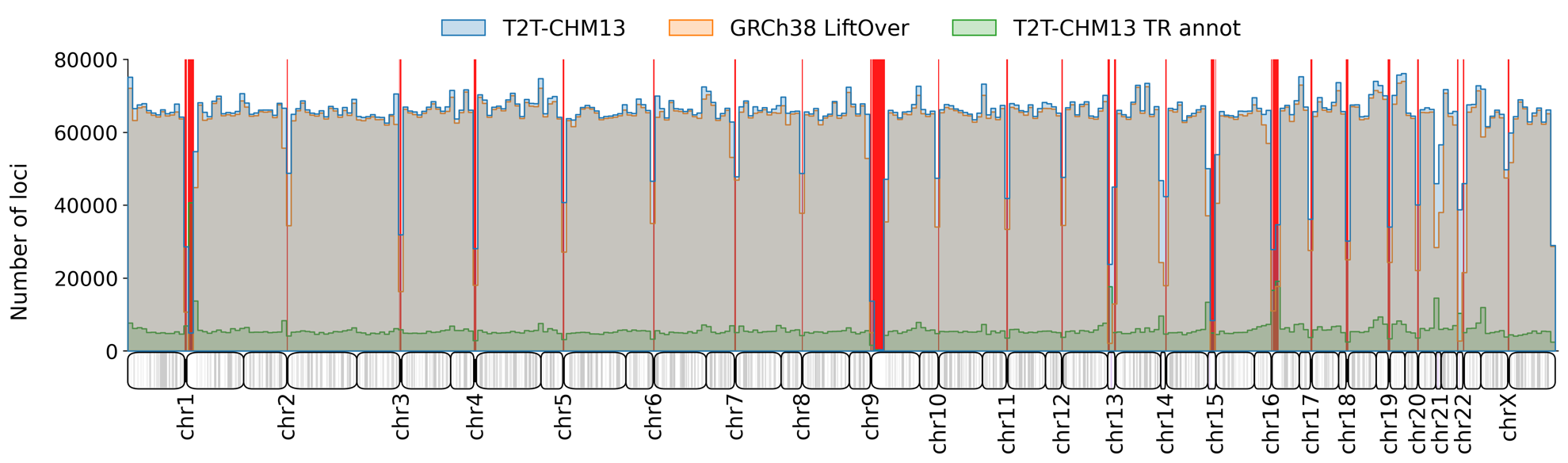

Supplementary Figure 3. Histogram showing the distribution of TR loci genotyped in the T2T-CHM13 catalog (blue) across the human genome (bin size = 1Mb). Distributions of loci from the GRCh38 catalog liftovered to T2T-CHM13 reference coordinates (orange) and the UCSC TR (Simple Repeats + RepeatFinder + Low Complexity) annotations (green) are also shown for comparison (overlapping blue and orange bars lead to grey colors). Red bars indicate T2T-CHM13 heavy repeat regions: regions >= 1Mb after merging TRs <= 100bp apart.

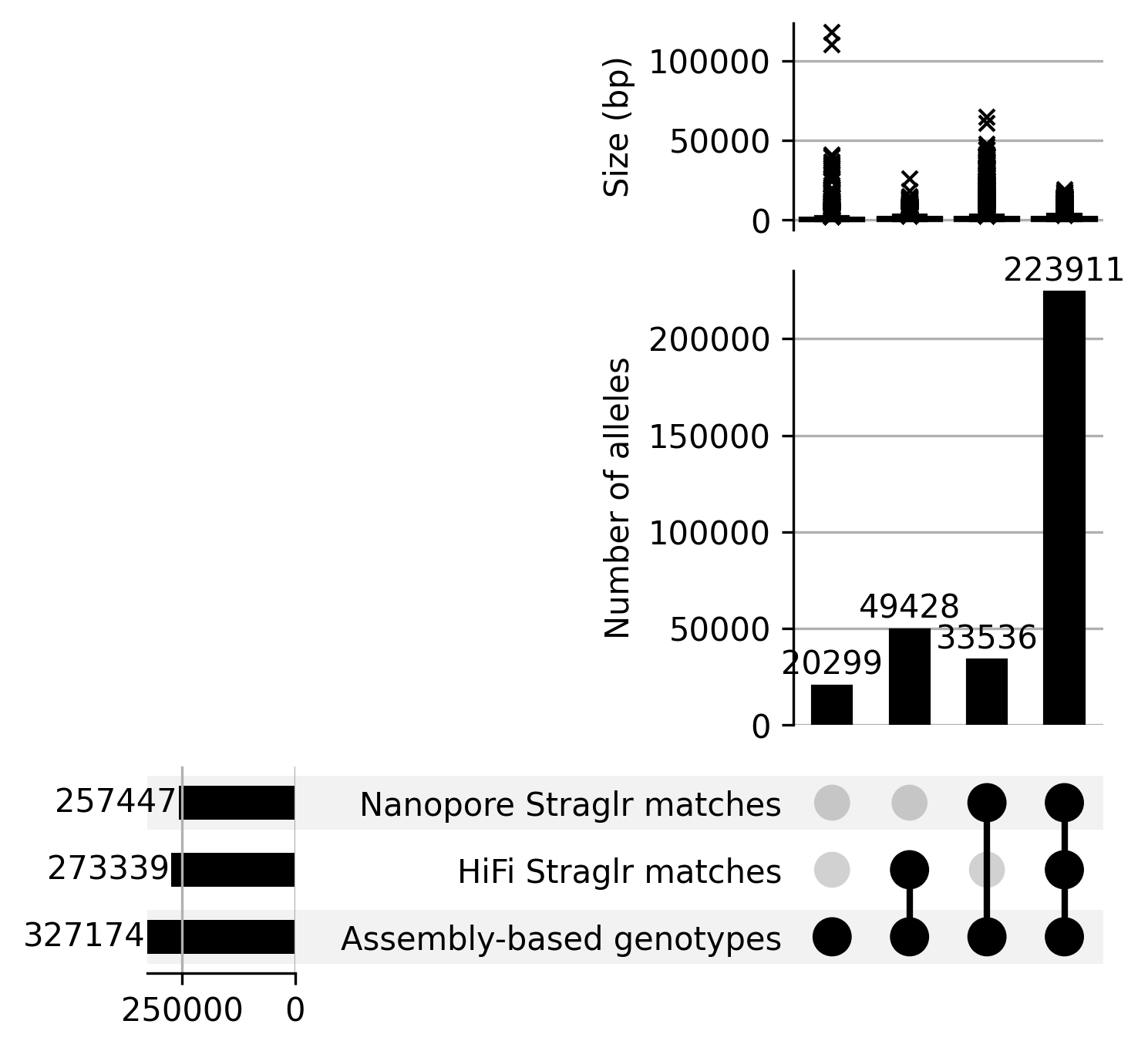

Supplementary Figure 4. Agreement between assembly-based and read-based genotypes. Overlaps (number of alleles) between TR genotypes in HPRC samples with Nanopore sequencing data reported by 1) Straglr on Nanopore sequencing reads; 2) Straglr on HiFi sequencing reads; and 3) assembly-based genotyping. (top) Number of alleles in each intersection; (bottom) Size (bp) distribution of alleles within each intersection below.

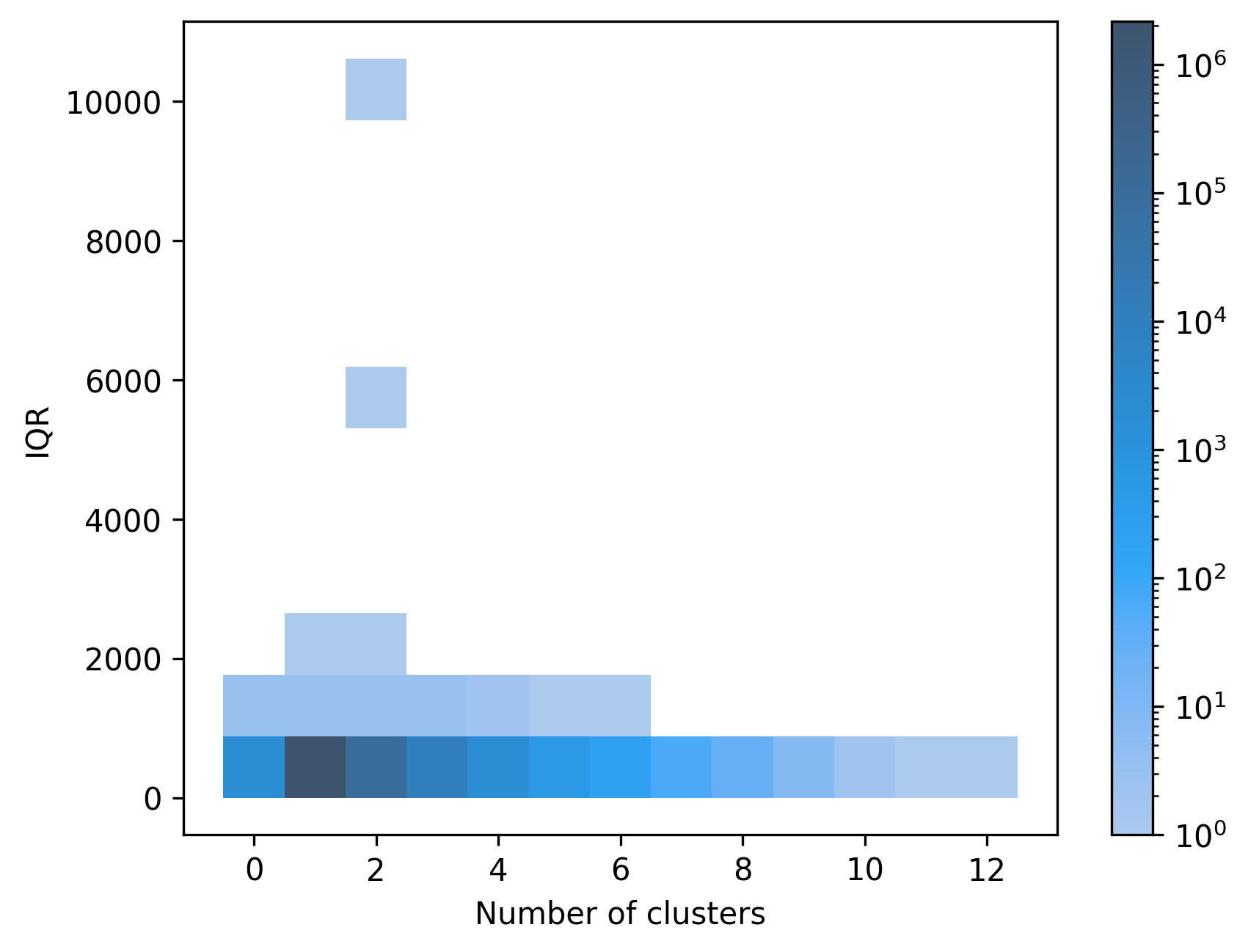

Supplementary Figure 5. Tallies of loci at different levels of size polymorphism measured by IQR and number of clusters. Number of loci (heatmap scale) with different combinations of IQR (Y-axis) and number of clusters based on repeat copies (X-axis).

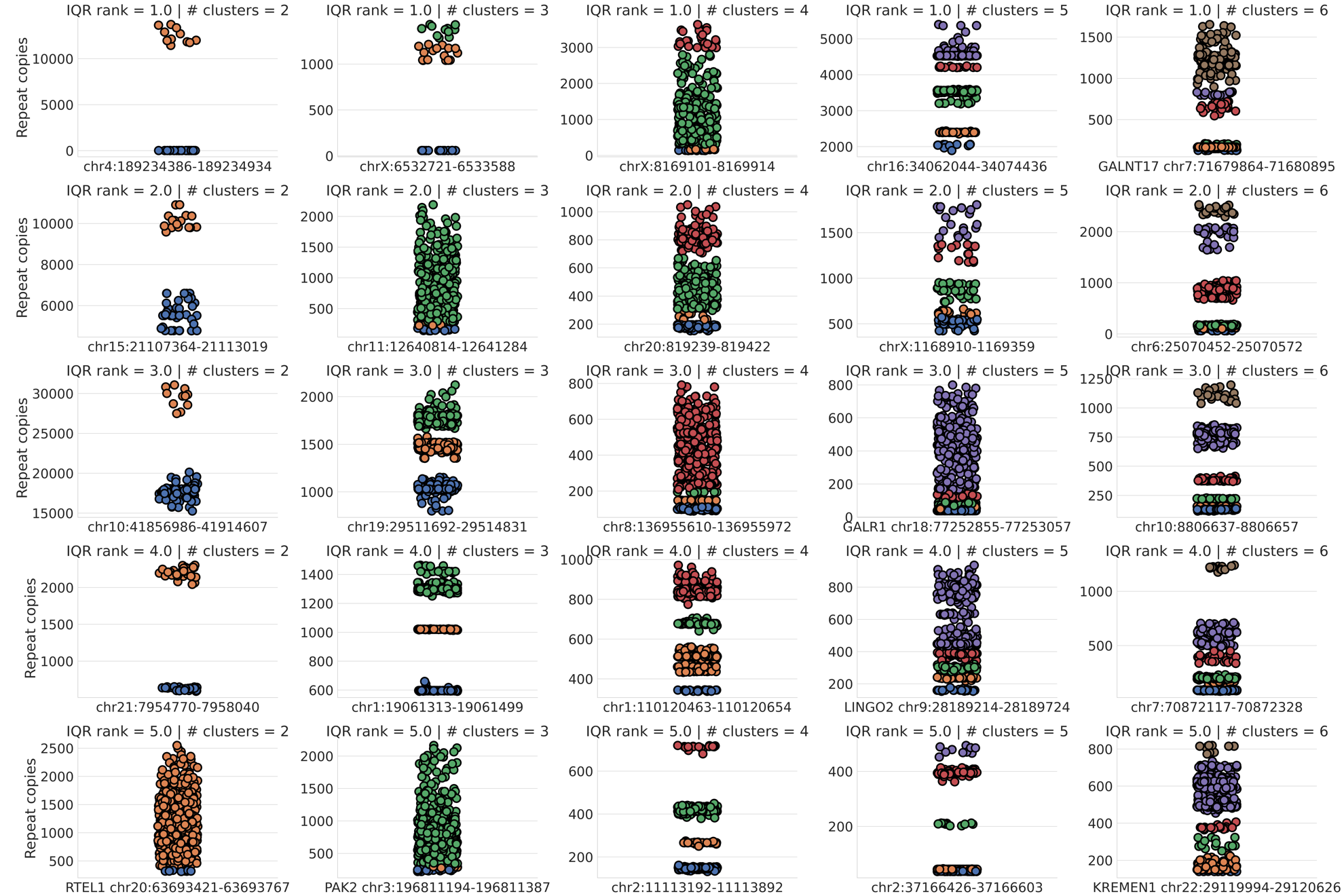

#### Supplementary Figure 6. Top cases of loci with high variance (IQR) in repeat copies and different levels of clustering.

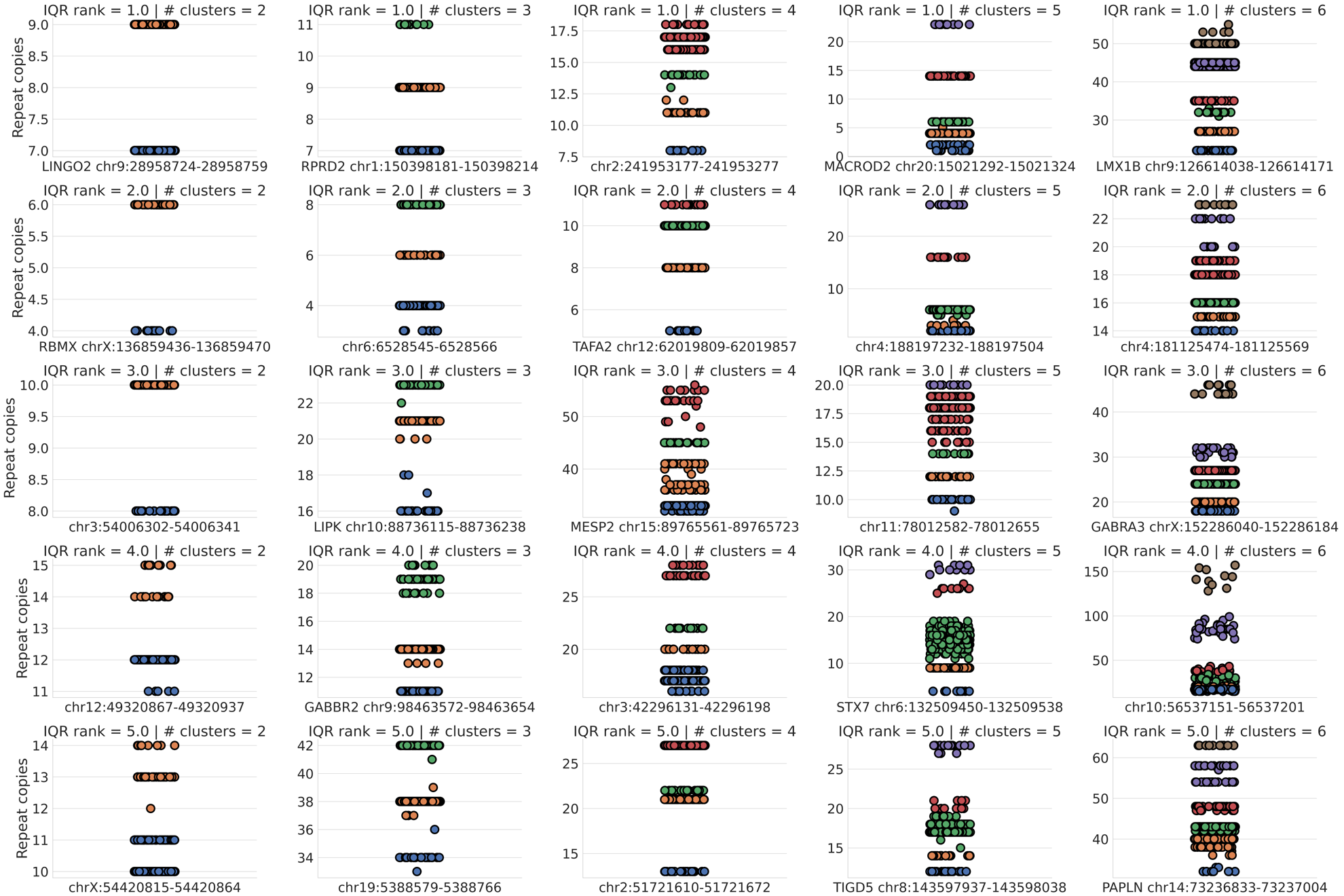

#### Supplementary Figure 7. Top cases of loci with low variance (IQR) in repeat copies and different levels of clusters.

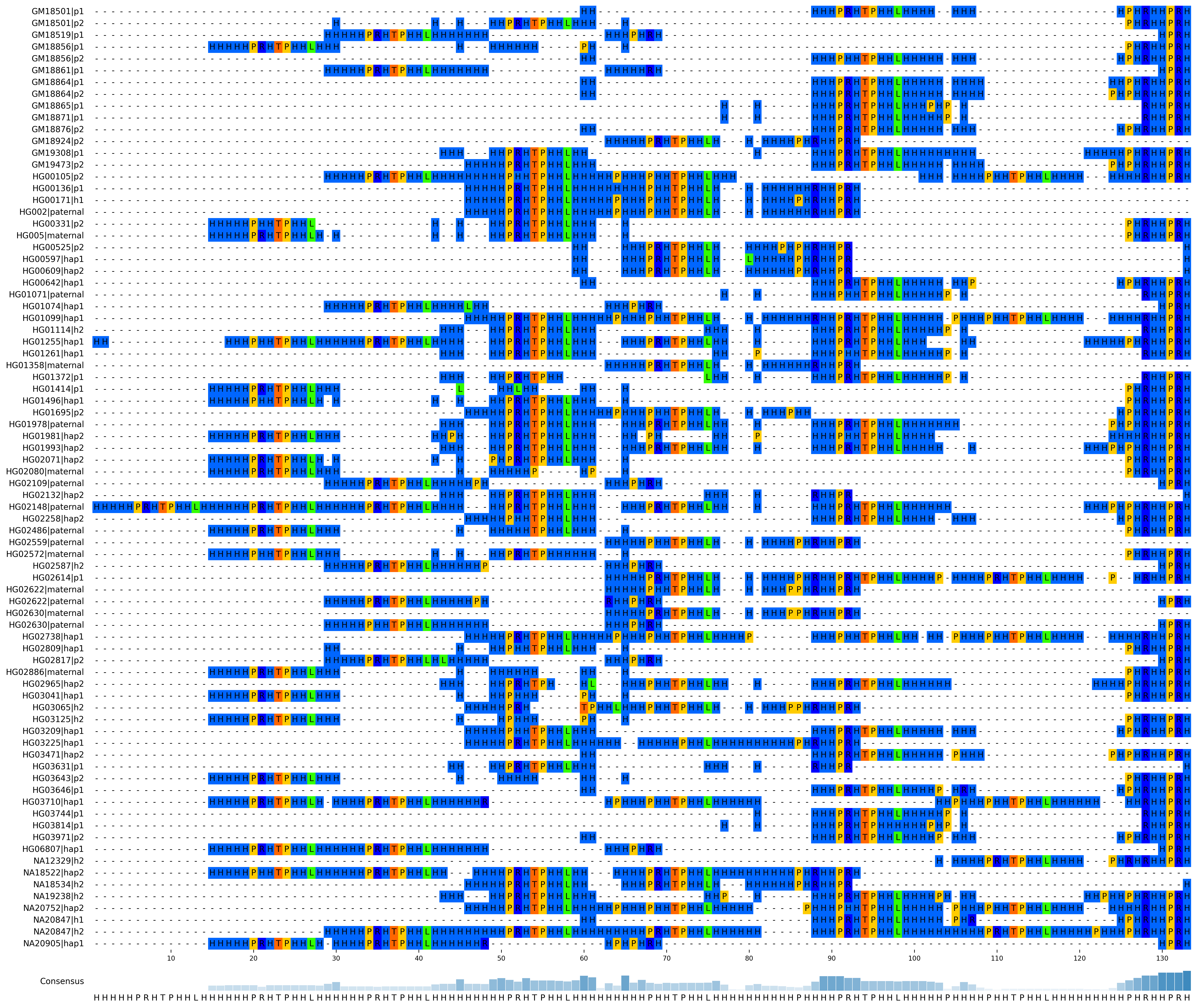

Supplementary Figure 8. Multiple sequence alignment of translated amino sequences of distinct assembled STRs at chr9:35,906,548-35,906,626 in exon 1 of *HRCT1*. Each sequence may be detected in multiple samples in our study cohort and only one representative (labeled by sample name and haplotype) was chosen for alignment.

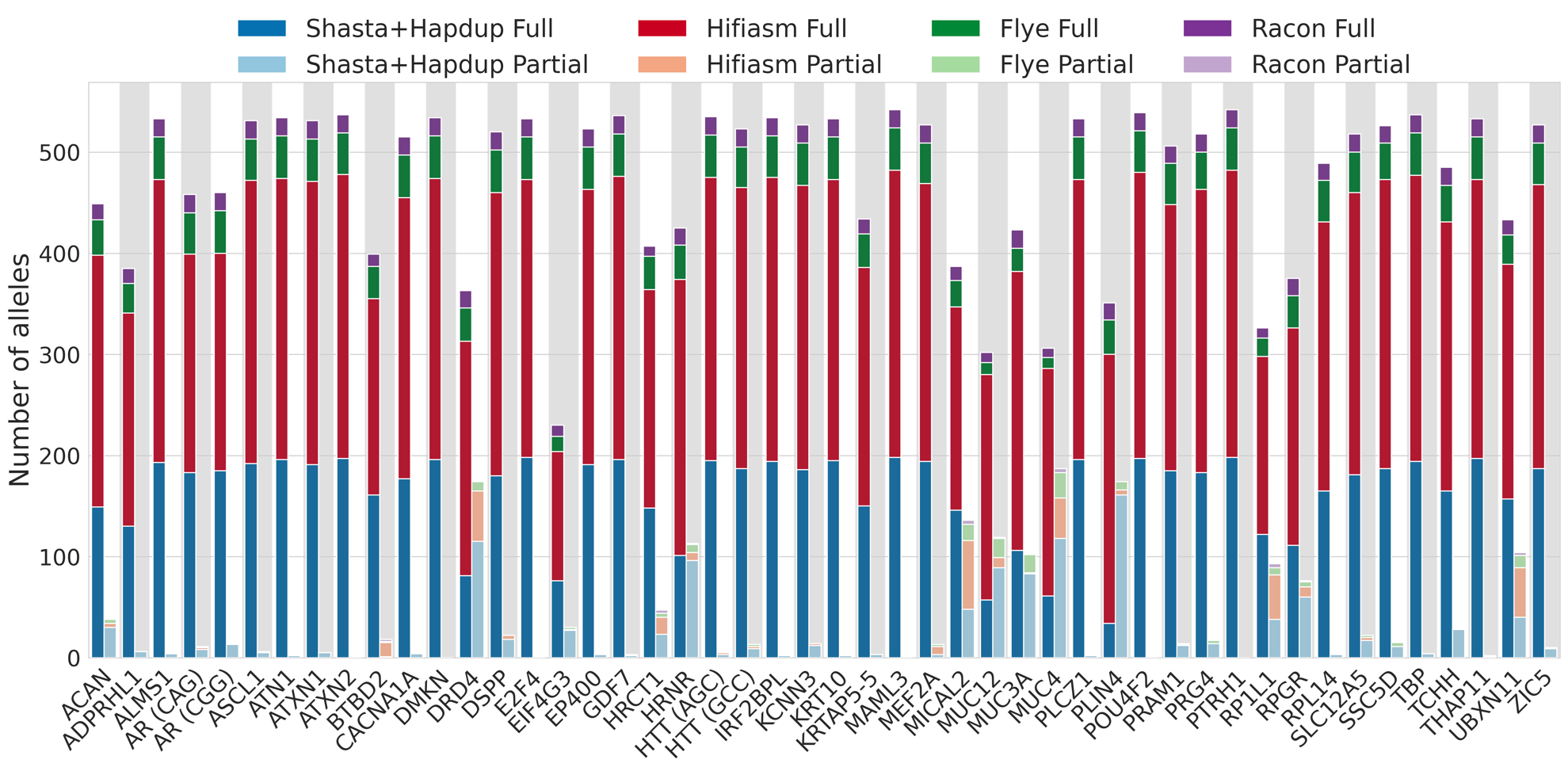

Supplementary Figure 9. Reconstruction of topmost polymorphic tandem repeats in coding sequences by assembler. “Full” (i.e. maintenance of opening reads frame, left stacked bars of each gene) and “Partial” (right stacked bars of each gene) labels assigned to reconstruction results determined by whether or not premature stop codons was introduced to translated amino sequence and hence leading to mismatch of downstream sequence.

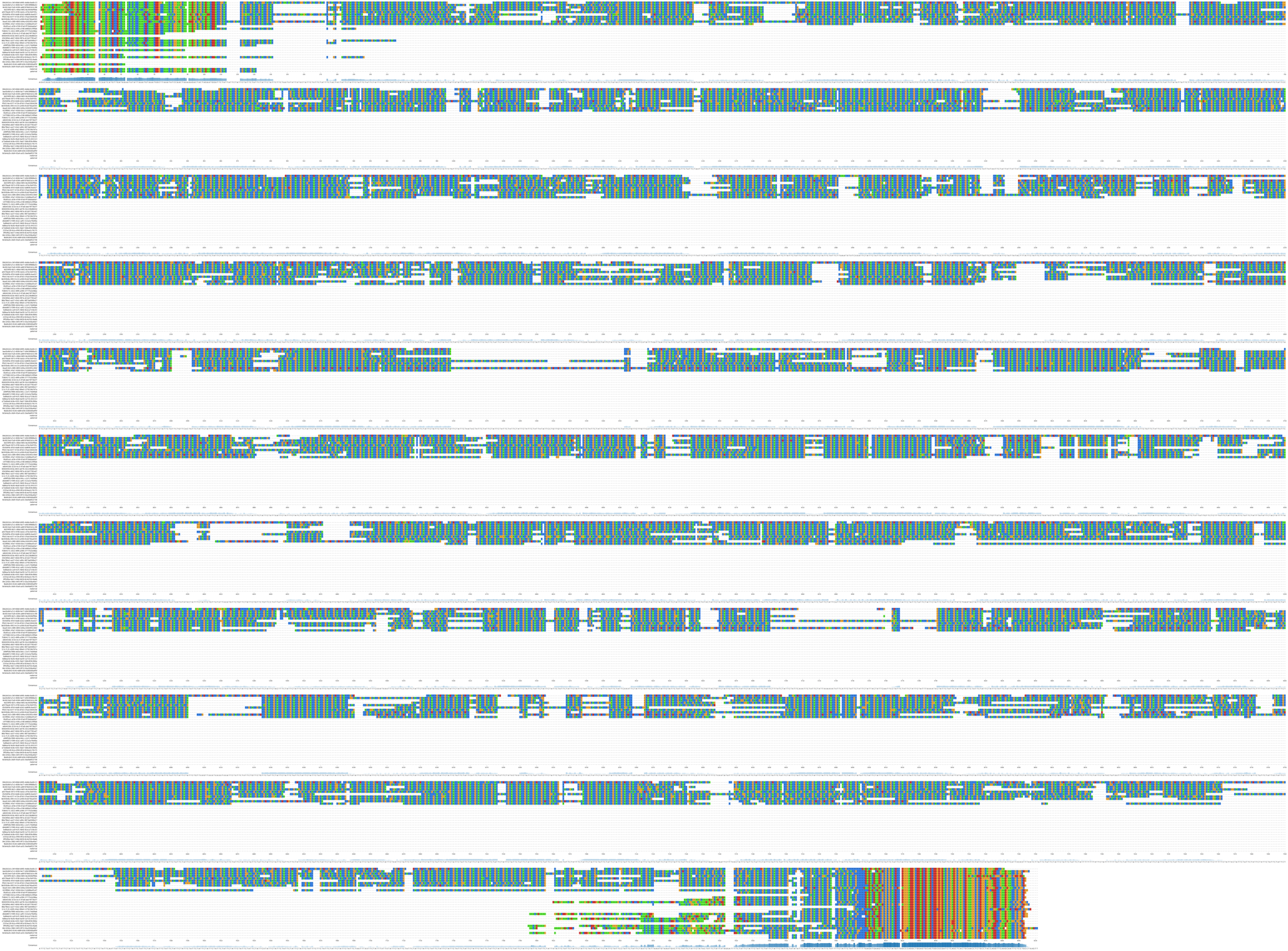

Supplementary Figure 10. Genotyping discrepancy at the ATTCT STR in ATXN10 in HG01123. Multiple sequence alignment of Nanopore reads (labeled by read names) supporting a ~6.2kb allele (top 10 rows), a 65 bp allele (middle 18 rows), and the parental alleles (last two rows, labeled “maternal” and “paternal”) assembled by trio Hifiasm in the HPRC assembly of HG01123. The ATTCT repeat tract is manifested as mostly blue (color of nucleotide T) with specks of green (color of nucleotide A) and orange (color of nucleotide C). 100 bp of flanking sequences are included as shown on both ends of the alignments.

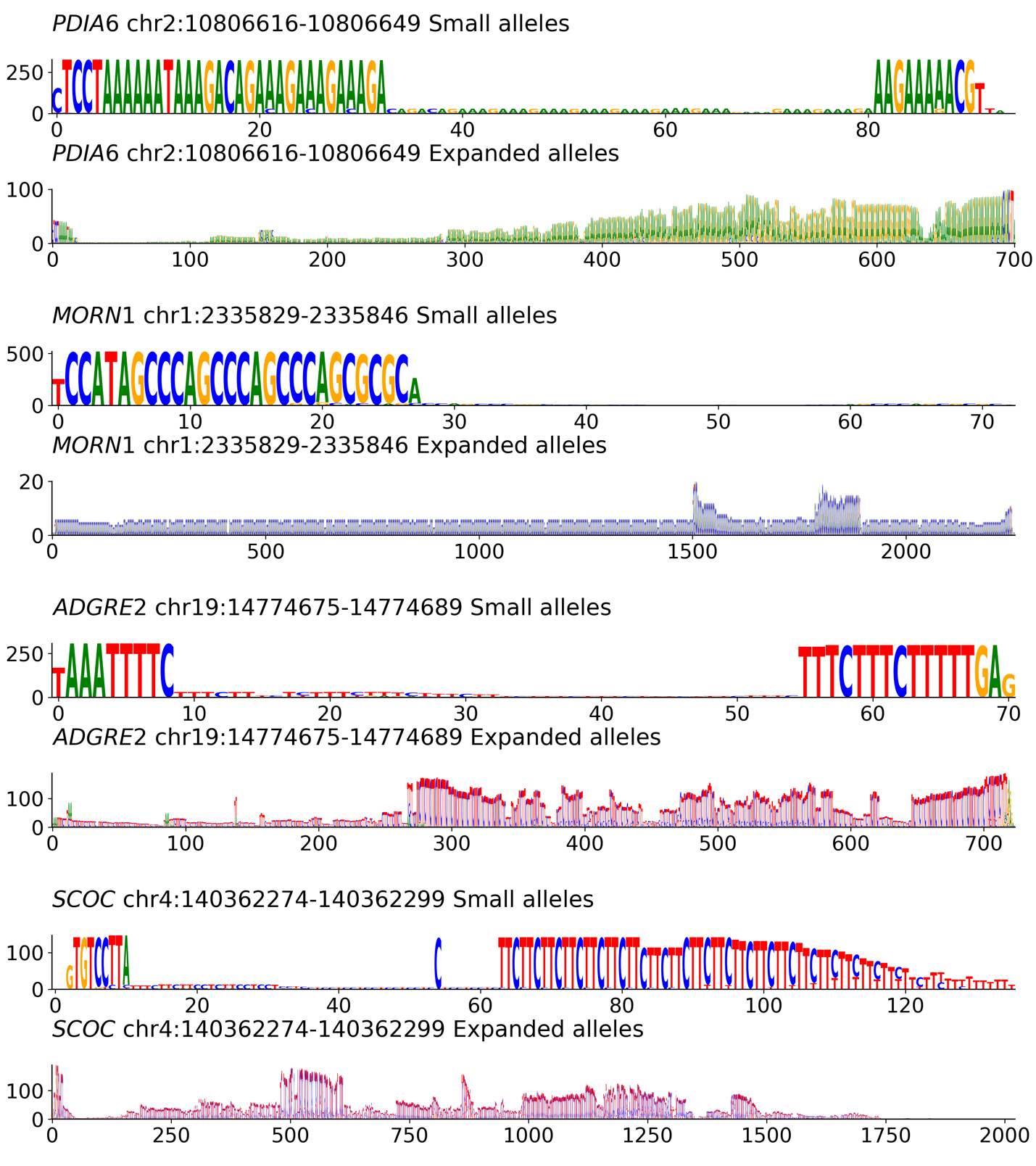

Supplementary Figure 11. Four unannotated STRs in genic loci with alleles larger than reference by at least 500 bp. Both small alleles with size close to reference (top) and expanded alleles (bottom) are shown as summarized in sequence logos. Y-axis indicates number of alleles; x-axis indicates sequence length (bp).

### Supplementary Tables

| Consortium | Location |
| --- | --- |
| HPRC | <https://s3-us-west-2.amazonaws.com/human-pangenomics/index.html?prefix=working/> |
| HGSVC2 | <https://ftp.1000genomes.ebi.ac.uk/vol1/ftp/data_collections/HGSVC2/release/v1.0/> |
| 1000G ONT | <https://s3.amazonaws.com/1000g-ont/index.html?prefix=FIRST_100_FREEZE/> |

#### Supplementary Table1. Online locations of the sequence data used in this study.

|  |  | GRCh38 | | T2T-CHM13 | |
| --- | --- | --- | --- | --- | --- |
| Sequencing Technology | Assembler | Edits | Novel loci | Edits | Novel loci |
| PacBio HiFi | Hifiasm (HPRC), Racon (HGSVC2) | 298 | 83 | 236 | 30 |
| PacBio CLR | Flye | 1,043 | 264 | 639 | 209 |
| Nanopore | Shasta+Hadup | 156 | 106 | 117 | 58 |

Supplementary Table 3. Average number of Straglr-based edits made to assembly-based genotypes per sample by sequencing technology and assembler. “Edits” = sum of corrections and missing haplotypes. “Novel loci” = loci from Straglr results that do not overlap loci from assembly-based genotypes.
